# Per-allele disease and complex trait effect sizes are predominantly African MAF-dependent in European populations

**DOI:** 10.64898/2025.12.31.25343290

**Authors:** Jordan Rossen, Benjamin J. Strober, Kangcheng Hou, Gaspard Kerner, Alkes L. Price

## Abstract

Understanding genetic architectures of disease is fundamental to partitioning heritability, polygenic risk prediction, and statistical fine-mapping. Genetic architectures of disease in European populations have been shown to depend on European minor allele frequency (MAF): SNPs with lower MAF have larger per-allele effects, due to the action of negative selection. However, we hypothesized that African MAF (defined using African-ancestry segments in African Americans), which is not distorted by the out-of-Africa bottleneck, might better predict per-allele effect sizes of common genetic variation in European populations; we note that common variants explaining most disease heritability are typically much older than the split between African and non-African populations. To demonstrate this, we first analyze the proportion of non-synonymous SNPs, which are strongly impacted by negative selection. The proportion of non-synonymous SNPs is much better predicted by African MAF than European MAF; a mixture of African MAF with weight *w*=0.95 (95% CI: (0.93, 0.96)) and European MAF with weight (1−*w*) is a more powerful predictor than either European MAF (*P*<10^-15^, 3.65x greater increase in log-likelihood relative to a null model without MAF dependence) or African MAF (*P*<10^-15^). Next, we consider the widely used 𝛼 model, in which per-allele GWAS effect size variance is proportional to [𝑝_𝐸_(1 − 𝑝_𝐸_)]^𝛼^, where *p_E_* is the European MAF. We propose a different model in which per-allele effect size variance is proportional to [𝑝_𝑚𝑖𝑥_(1 − 𝑝_𝑚𝑖𝑥_)]^𝛼𝑚𝑖𝑥^, where *p_mix_*=*w*\**p_A_*+(1−*w*)\**p_E_,* and *p_A_* is the African MAF. We fit the *α_mix_* model by extending the baseline-LD model used in S-LDSC to include a grid of bivariate African and European MAF bins and identifying values of *w* and *α_mix_* that best fit mean effect size variance estimates from S-LDSC across bivariate MAF bins. We demonstrate that our approach provides conservative estimates of *w* in simulations. We applied this approach to summary statistics for 50 diseases/complex traits in European populations (average *N*=483K) and estimated best-fit parameters of *w*=0.96 (95% CI: (0.76, 1.16)) and *α_mix_*=−0.34 (95% CI: (-0.67, -0.02)), attaining a far better fit than the standard 𝛼 model using *p_E_* only (*P<*10^-15^, 4.53x greater decrease in mean-squared error relative to a null model without MAF dependence). We conclude that per-allele disease and complex trait effect sizes are predominantly African MAF-dependent in European populations.

## Introduction

Understanding genetic architectures is fundamental to partitioning heritability^1–5^, polygenic risk prediction^6–11^, and statistical fine-mapping^12–16^. Recent studies have shown that genetic variants with lower minor allele frequency (MAF) tend to have larger per-allele effects on diseases and complex traits, due to negative selection against disease-associated variants that impact fitness^17–20^. This relationship is frequently modeled using the 𝛼 model^4,9,15,17–38^, in which the per-allele effect size variance of a SNP in a GWAS population (e.g. Europeans) is a function of its MAF in the GWAS population. However, limited attention has been paid to the possibility that the MAF in a different population (e.g. Africans) might be informative for GWAS architectures (see ref.^28,34,39^ and Discussion).

We hypothesized that (unadmixed) African MAF might better predict per-allele effect sizes in European populations, as it is not distorted by the out-of-Africa bottleneck and thus more accurately reflects the consequences of negative selection; we note that common variants explaining most disease heritability are typically older than the split between African and non-African populations^40,41^. First, we demonstrate that the proportion of SNPs that are non-synonymous is better predicted by African MAF than European MAF, relying on non-synonymous variant type as a partial proxy for variant effect size. Second, we analyze European GWAS summary statistics using a model that incorporates both African MAF and European MAF to demonstrate that estimated per-allele effect variance is more heavily influenced by African MAF than European MAF.

## Results

### Overview of methods: defining the *α_mix_* model

We propose the *α_mix_* model for quantifying the MAF-dependence of per-allele effects on diseases/traits. We model per-allele effect size variance as a function of a linear combination of (unadmixed) African and European MAF, instead of MAF in a single ancestry as in the widely used 𝛼 model^4,9,17–38^.

In detail, the *α_mix_* model relates per-allele effect size variance to unadmixed African (*p_A_*) and European (*p_E_*) MAF:

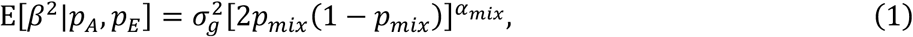

where 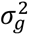 is a variance parameter, *p_mix_*=*w*\**p_A_*+(1−*w*)\**p_E_*, *w* specifies the relative contributions of *p_A_* and *p_E_*, and *α_mix_* specifies the level of MAF dependence.

In the *α_mix_* model, we define African MAF (*p_A_*) using whole-genome sequencing data from All of Us^42^ (*N*=46,672 African Americans, correcting for European admixture by extracting local genomic regions with two African haplotypes identified using RFMix2^43^ (Methods)) and 1000 Genomes^44^ (*N*=107 Yoruba), and define European MAF using whole-genome sequencing data from 1000 Genomes^44^ (*N*=489 Europeans); we did not use All of Us to define *p_E_* to ensure consistency with the baseline-LD (v2.2) model^3,5^. In analyses of non-synonymous variation, we define African and European MAF using whole-genome sequencing data from All of Us (*N*=46,672 African Americans and *N*=70,346 European Americans). We note that Africa harbors a diverse set of populations^45–47^, but we focused on the unadmixed African component of African Americans, due to (i) the large available sample size and (ii) its known genetic similarity (*F*_ST_ < 0.001) to the Yoruba population^48^, which is widely studied and has not been subject to recent extreme bottlenecks^44^.

### Overview of methods: fitting the *α_mix_* model

We fit the *α_mix_* model by stratifying SNPs into bins based on African and European MAF, estimating the mean per-allele effect size variance of each bin using S-LDSC^3,5^ and estimating *α_mix_* model parameters that best fit the S-LDSC estimates; this procedure is similar to the approach used in ref.^18^ to fit the 𝛼 model.

In detail, we estimate *α_mix_* via a generalized method of moments estimator. First, we define a grid of bivariate MAF bins, where each bivariate MAF bin is defined by an African MAF bin and a European MAF bin. Second, we estimate the mean effect size variance of each bivariate MAF bin using S-LDSC^3^ with an extension of the baseline-LD (v2.2) model^5^ designed to capture African MAF-dependence (Methods). Finally, we use a numerical optimization procedure to estimate values of *α_mix_*, 𝑤, and 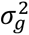 that minimize the mean squared error (summed across bivariate bins and weighted by the number of SNPs in each bin) between the mean effect size variance estimated by S-LDSC and E[𝛽^2^|𝑃_𝐴_, 𝑃_𝐸_] as computed using Equation 1 (averaging across values of *p_mix_\** (1−*p_mix_*) in each bivariate MAF bin). We fit Equation 1 using SNP bins (rather than individual SNPs) for two reasons. First, the expected effect size variance estimated by S-LDSC is often negative for individual SNPs, but is less frequently negative for bivariate MAF bins. Second, *p_mix_\** (1-*p_mix_*) for individual SNPs can approach 0 when *w* is close to 1, but does not approach 0 when averaged across bivariate MAF bins. In analyses across diseases/traits, we estimate *w* and *α_mix_*jointly while estimating 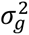 separately for each trait; we also include analyses in which we estimate 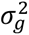, *w*, and 𝛼 separately for each trait. We have publicly released custom code used to estimate *α_mix_* and perform all analyses described in this manuscript (Code Availability).

We analyze summary association statistics for 50 approximately independent, heritable diseases/traits in European ancestry populations (29 UK Biobank and 21 other diseases/traits; average *N*=483K) (Methods and Supplementary Table 1). We have publicly released summary association statistics and S-LDSC output for each disease/trait (Data Availability).

We complement our analyses of disease/trait heritability by analyzing how the proportion of non-synonymous SNPs varies with African MAF and European MAF. We use the proportion of non-synonymous SNPs as a partial proxy for per-allele effect size variance, as non-synonymous variants are more strongly impacted by negative selection^18,49–53^, and are highly enriched for heritability^11,18,53^ and fine-mapped causal effects^14,54,55^. We model the probability that a SNP is non-synonymous as a function of *p_mix_* via the following logistic regression model:

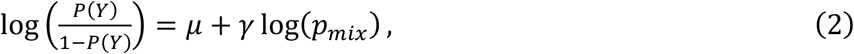

where *Y*=1 if the SNP is non-synonymous and 0 otherwise and *p_mix_*=*w*\**p_A_*+(*1*-*w*)\**p_E_* as above (Methods). To estimate *w*, we compute a profile likelihood, treating 𝜇 and 𝛾 as nuisance parameters, and use a numerical optimization method to maximize the profile likelihood with respect to *w* (Methods).

### The proportion of non-synonymous SNPs is predominantly African MAF-dependent in European and African ancestry populations

We hypothesized that the probability that a SNP is non-synonymous might be better predicted by its African MAF than its European MAF, because African MAF is not distorted by the out-of-Africa bottleneck. To evaluate this hypothesis, we calculated the proportion of SNPs that are non-synonymous (PNS), stratified by bivariate African and European MAF bins (Figure 1a, Supplementary Figures 1-2 and Supplementary Table 2). In general, decreasing African MAF within a European MAF bin corresponds to a substantial decrease in PNS, whereas decreasing European MAF within an African MAF bin corresponds to a less substantial decrease in PNS. To quantify this finding, we fit logistic regression models for non-synonymous status as a function of log(*p_A_*) stratified by *p_E_* (Figure 1b and Supplementary Table 3-4), and as a function of log(*p_E_*) stratified by *p_A_* (Figure 1c, Supplementary Figure 3, and Supplementary Table 5-6). We observed consistently strong effects for *p_A_* within each *p_E_* bin (𝛾=−0.10, −0.13, and −0.13, for *p_E_ ≥* 0.05, 0.005 ≤ *p_E_* < 0.05, and *p_E_* < 0.005, respectively; *P<*10^-15^ for each bin), but generally weaker effects for *p_E_* within each *p_A_* bin (𝛾=−0.028, −0.047, and −0.10, for *p_A_ ≥* 0.05, 0.005 ≤ *p_A_* < 0.05, and *p_A_* < 0.005, respectively; *P<*10^-15^ for each bin); the relatively stronger effect for *p_A_* < 0.005 may reflect recent selection on variants that arose in Europeans after the out-of-Africa bottleneck^52^, including variants that arose via Neanderthal introgression^56–58^. This supports the hypothesis that African MAF is highly predictive of PNS, even after accounting for European MAF.

**Figure 1:**
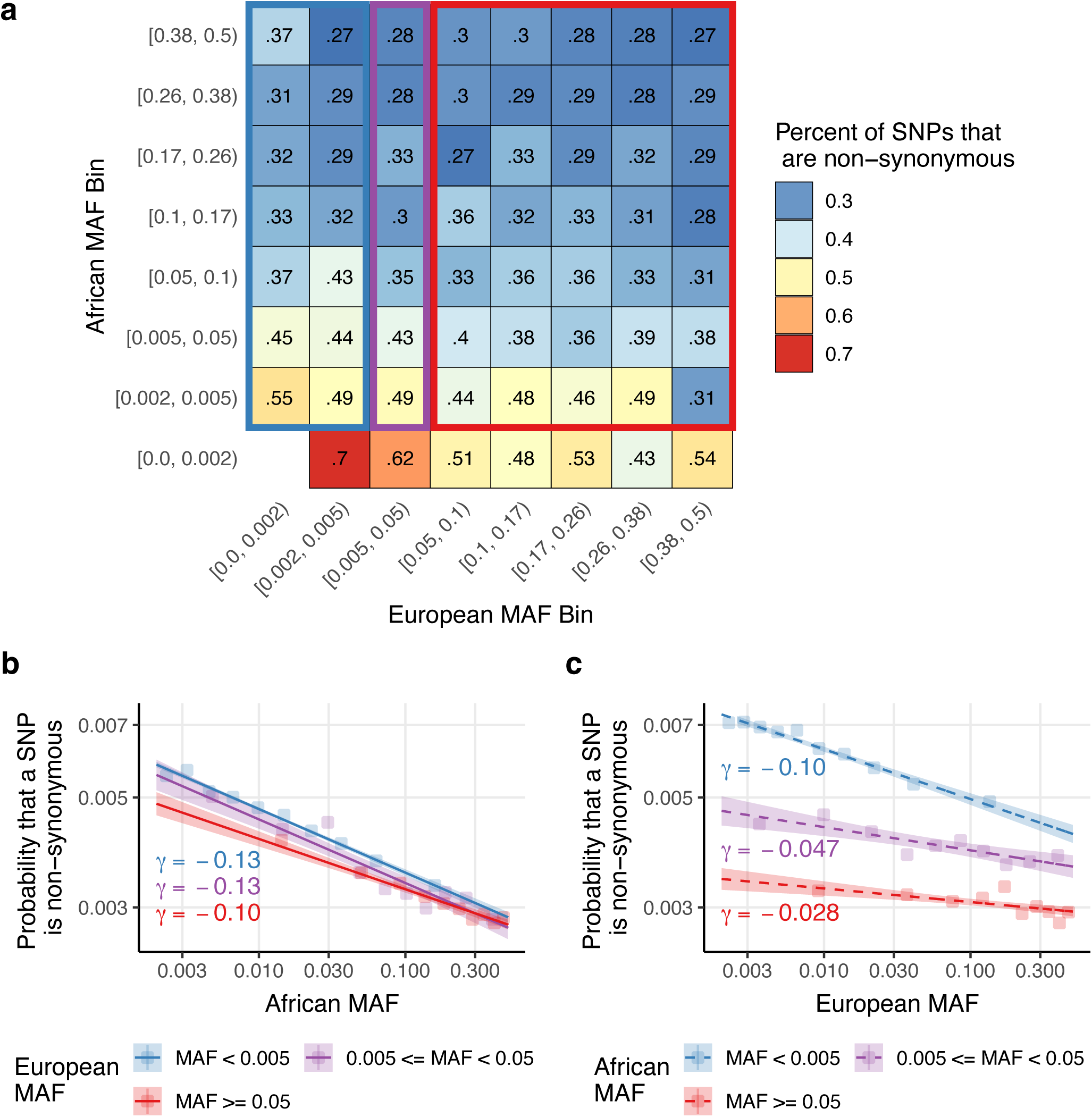
The proportion of non-synonymous SNPs as function of unadmixed African and European minor allele frequencies: We report **(a)** the percentage of SNPs that are non-synonymous in bivariate bins defined by unadmixed African MAF (rows) and European MAF (columns), **(b)** the probability that a SNP is non-synonymous as a function of African MAF after stratifying by European MAF categories (for SNPs with *p_A_* ≥ 0.002) as predicted by logistic regression, and **(c)** the probability that a SNP is non-synonymous as a function of European MAF after stratifying by African MAF categories (for SNPs with *p_E_* ≥ 0.002) as predicted by logistic regression. The blue, purple, and red rectangles in (a) correspond to the SNPs included in the blue, purple, and red lines in (b). The choice of MAF bin boundaries is discussed in Methods. Estimated regression coefficients for African MAF within European MAF categories are given in the bottom left of (b). Estimated regression coefficients for European MAF within African MAF categories are given below each line in (c). Shaded regions in (b) and (c) denote 95% confidence intervals on the mean estimate. Logistic regression fits with one regression per plot are reported in Supplementary Figure 3. Numerical results, including standard errors for each bivariate bin in (a) and each bin in (b) and (c), are reported in Supplementary Tables 2-6.

To directly evaluate whether African MAF are more predictive of PNS than European MAF, we fit a logistic regression model for non-synonymous status as a function of log(*p_mix_*) (where *p_mix_*=*w*\**p_A_*+(1-*w*)\**p_E_*) (see Equation 2) and compared different values of *w* via a profile likelihood approach, treating 𝜇 and 𝛾 as nuisance parameters. We restricted to SNPs with *p_A_* ≥ 0.002 and/or *p_E_* ≥ 0.002 (MAF thresholds are discussed in Methods). The maximum likelihood estimate (MLE) of *w* was 0.95 (95% CI: (0.93, 0.96)) (Figure 2a and Supplementary Table 7), indicating that PNS is predominantly dependent on African MAF, but European MAF provides some additional information (*P*<10^-15^ for comparisons to European MAF-only or African MAF-only models).

**Figure 2:**
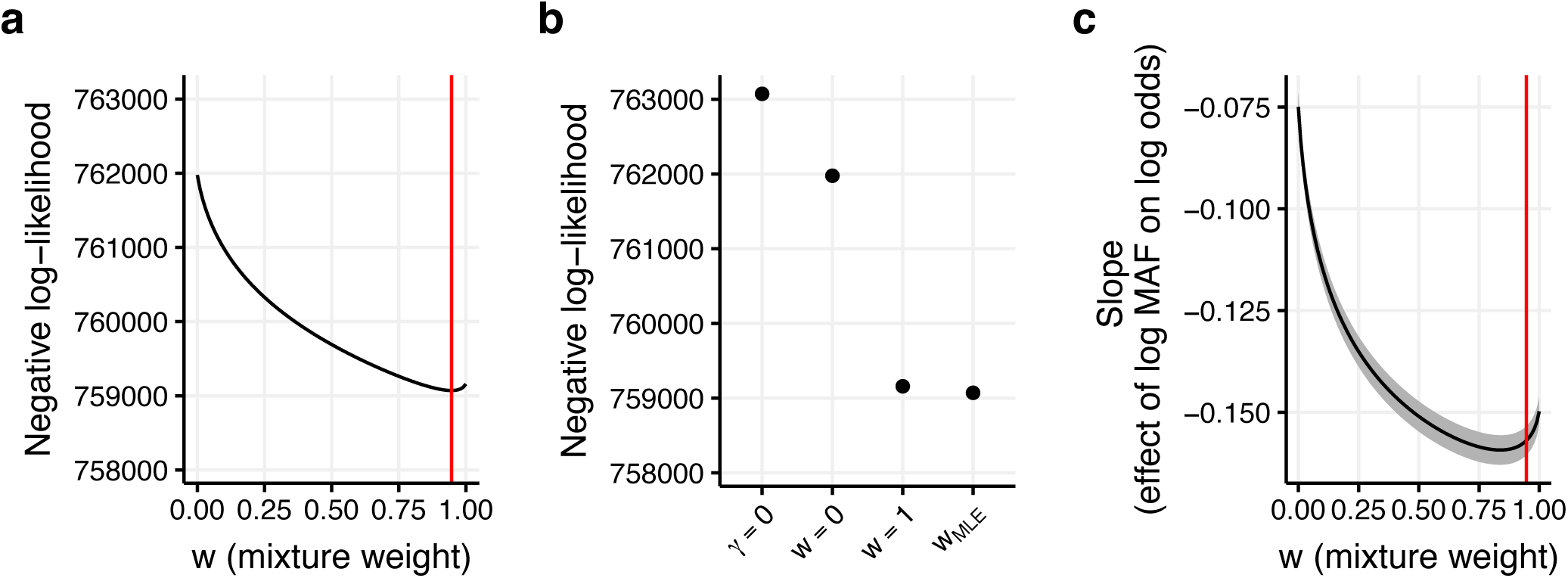
Best-fit parameters for a logistic regression of non-synonymous status on log(*p_mix_*). We report **(a)** the negative profile log-likelihood for various models, including a model without MAF dependence (𝛾=0), a model with European MAF dependence (*w*=0), a model with African MAF dependence (*w*=1) and the mixture model from equation 2 (*w_MLE_*), **(b)** the negative profile log-likelihood with respect to *w*, treating the regression intercept and slope as nuisance parameters, and **(c)** the regression slope (𝛾) for values of *w* between 0 and 1. Red lines in (b) and (c) denote the MLE estimate of *w*. The shaded region in (c) denotes 95% confidence intervals for 𝛾. Numerical results are reported in Supplementary Table 7.

Compared to a null model with no MAF dependence (𝛾=0), the *p_mix_* model attained a 3.65x larger increase in log-likelihood than the European MAF-only model (*w*=0) and a 1.02x larger increase in log-likelihood than the African MAF-only model (*w*=1) (Figure 2b and Supplementary Table 7).

Interestingly, the value of *w* that produced the largest absolute 𝛾 (*w*=0.84) (Figure 2c and Supplementary Table 7) was smaller than the MLE value (*w*=0.95), because varying *w* changes the variance of log(*p_mix_*); this discrepancy became negligible when log(*p_mix_*) was standardized to have unit variance (Supplementary Figure 4).

We performed 5 secondary analyses. First, we re-estimated the effects of conditional effects of *p_A_* (stratifying by *p_E_*) and *p_E_* (stratifying by *p_A_*) on PNS while restricting to SNPs with both *p_A_* ≥ 0.002 and *p_E_* ≥ 0.002. Results were similar to Fig. 1b-c (Supplementary Figure 5). Second, we compared the marginal effect of *p_A_* on PNS to the marginal effect of *p_E_* on PNS using logistic regression for five sets of SNPs: SNPs with *p_A_* ≥ 0.002 and/or *p_E_* ≥ 0.002, SNPs with *p_A_* ≥ 0.002 and *p_E_* ≥ 0.002, SNPs with *p_A_* ≥ 0.002, SNPs with *p_E_* ≥ 0.002, and SNPs with *p_A_* ≥ 0.05 and *p_E_* ≥ 0.05. African MAF provided a better fit for all five SNP sets (*P <* 10^-15^ for all tests, except *P* = 0.01 for SNPs with MAF ≥ 0.05 in both ancestries) (Supplementary Figure 6). Third, we compared the marginal effect of *p_A_* on PNS for all SNPs with *p_A_* ≥ 0.002 to the marginal effect of *p_E_* on PNS for all SNPs with *p_E_* ≥ 0.02 (Supplementary Figure 7). Surprisingly, the *p_E_* model attained a higher McFaddens pseudo-*r*^2^ and higher absolute slope than the *p_A_* model (McFadden’s pseudo-*r*^2^=0.007 for *p_E_* vs. 0.004 for *p_A_*, 𝛾=−0.17 for *p_E_* vs. −0.13 for *p_A_*, *P* < 10^-15^ for pseudo-*r^2^* and 𝛾 differences); however, we note that this comparison varies both the set of SNPs evaluated and the explanatory variable, and thus does not directly indicate whether *p_A_* or *p_E_* better predicts PNS for any fixed set of SNPs. We hypothesize that the observed difference in *r^2^* is because PNS is harder to predict for African-specific SNPs than for European-specific SNPs. Fourth, we refit the logistic regression on log(*p_mix_*) using 4 alternative SNP sets: SNPs with *p_A_* ≥ 0.002 and *p_E_* ≥ 0.002, SNPs with *p_A_* ≥ 0.002, SNPs with *p_E_* ≥ 0.002, and SNPs with *p_A_* ≥ 0.05 and *p_E_* ≥ 0.05. In each case, the MLE estimate for *w* was between 0.76-0.95, with tight confidence intervals (Supplementary Figure 8). Fifth, we refit the logistic regression on log(*p_mix_*) model while changing *p_A_* to max(0.05, *p_A_*) and *p_E_* to max(0.05, *p_E_*) (Supplementary Figure 9), rather than 0.002 as in the main analyses. The MLE for *w* was 0.80, similar to the estimate obtained when restricting to SNPs with *p_A_* ≥ 0.05 and *p_E_* ≥ 0.05.

### Per-allele disease and complex trait effect sizes are predominantly African MAF-dependent in European populations

We hypothesized that per-allele disease and complex trait effect sizes (e.g. in European populations, for which data is available in large sample size) might be better predicted by African MAF than by European MAF, because African MAF is not distorted by the out-of-Africa bottleneck. To evaluate this hypothesis, we used S-LDSC^3,5^ to estimate mean per-allele effect size variances across bivariate MAF bins (7 African MAF bins ranging from 0 to 0.50 x 5 European MAF bins ranging from 0.05 to 0.50), scaled to a mean value of 1 across SNPs and meta-analyzed across 50 diseases/traits (average *N*=483K; Supplementary Table 1) (Methods); we excluded SNPs with European MAF less than 0.05 because S-LDSC typically estimates SNP-heritability only for common SNPs^3,5^. Results are reported in Figure 3 and Supplementary Tables 8-9. We determined that per-allele effect size variance exhibited a much stronger dependence on African MAF conditional on European MAF (1.1x-2.7x ratio within each of 5 common *p_E_* columns of Figure 3a, restricted to common *p_A_* rows; solid red line in Figure 3b) than European MAF conditional on African MAF (0.6x-1.4x ratio within each of 5 common *p_A_* rows of Figure 3a; dashed red line in Figure 3b). The dependence on African MAF was much stronger without restricting to SNPs with *p_E_* > 0.05 (1.8x-6.7x ratio; 5 common *p_E_* columns of Figure 3a); the dependence on European MAF was stronger for SNPs that are rare in Africans (dashed blue line in Figure 3b), possibly reflecting recent selection on variants that arose in Europeans after the out-of-Africa bottleneck^52^, including variants that arose via Neanderthal introgression^56–58^. These findings support the hypothesis that African MAF is highly predictive of per-allele effect size variance, even after accounting for European MAF.

**Figure 3:**
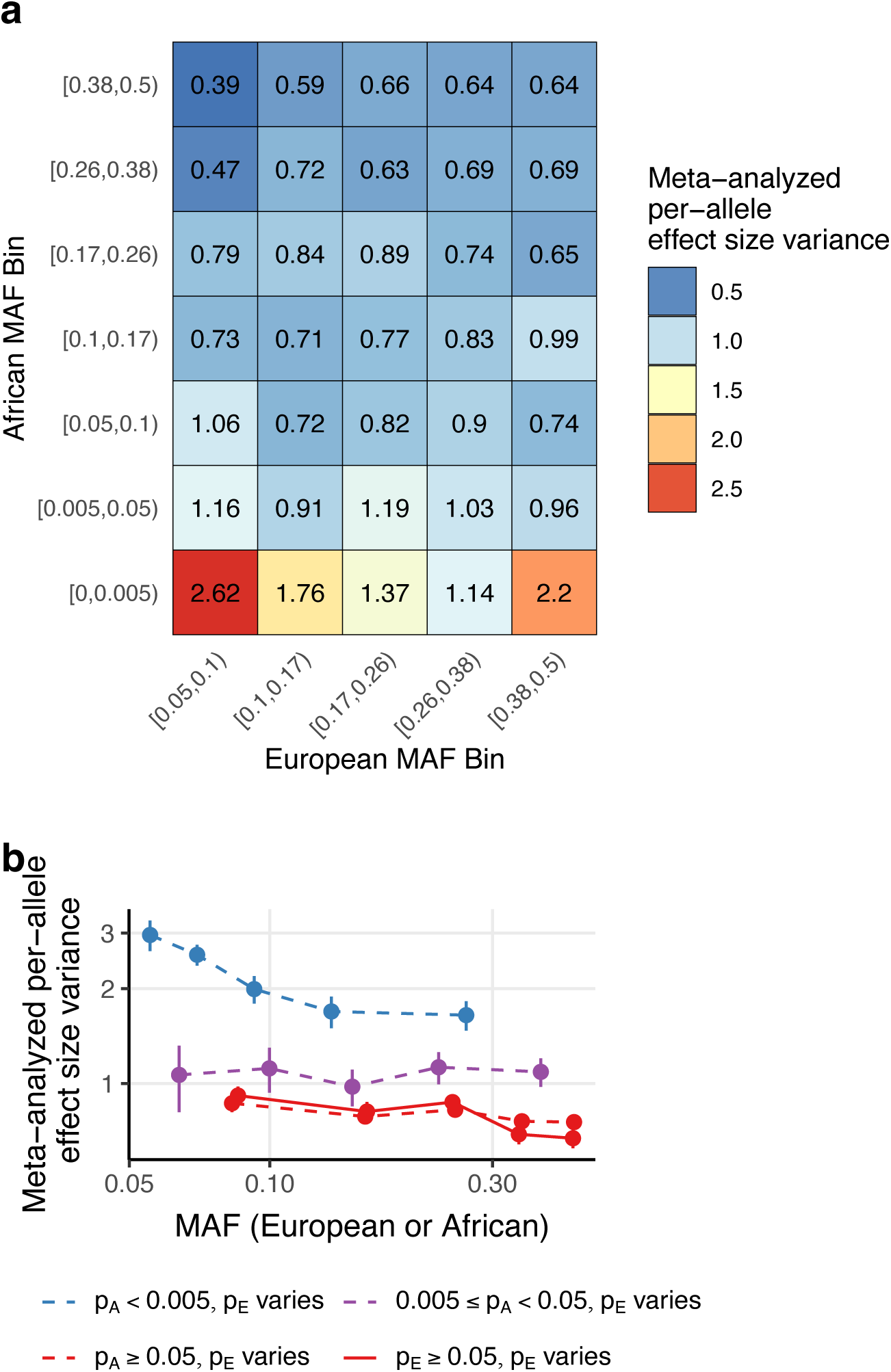
Per-allele effect size variance across 50 diseases/traits as a function of African and European minor allele frequencies. We report meta-analyzed scaled mean effect size variance as a function of **(a)** bivariate MAF bins defined by 7 African and 5 European MAF bins, and **(b)** African (solid line) and European (dashed lines) MAF quintiles, stratified by European and African MAF categories, respectively. The choice of MAF bin boundaries in (a) is discussed in Methods. Error bars in (b) denote 95% confidence intervals; error bars are smaller than dot size in some cases. Numerical results, including standard errors for each bivariate bin in (a) and each bin in (b), are reported in Supplementary Table 8-9.

To directly evaluate whether African MAF are more predictive of per-allele effect size variance than European MAF, we fit the *α_mix_* model (Equation 1 and Methods) across the 50 diseases/traits, using a single *w* parameter and a single *α_mix_* parameter. Results are reported in Figure 4 and Supplementary Table 10. Best-fit estimates were *w*=0.96 (95% CI: (0.76, 1.16)) and *α_mix_*=−0.34 (95% CI: (−0.67,−0.03)), indicating that per-allele effect size variance is best predicted by placing much greater weight on African MAF than on European MAF (*P*<10^-15^ vs. null hypothesis of *w*=0 (𝛼 model); *P*=9*10^-6^ vs. null hypothesis of *w*=0.5) (Figure 4a); we failed to reject the null hypothesis that per-allele effect size variance depends on African MAF alone (*P=*0.71 vs. null hypothesis of *w*=1). Relative to a null model with no MAF dependence (*α_mix_* = 0), *α_mix_*attained a 4.53x greater decrease in weighted MSE than a European MAF-only model (*w*=0) and a 1.03x greater decrease in weighted MSE than an African MAF-only model (*w*=1) (Figure 4b). The relationship between *w* and the MSE of the *α_mix_* model was remarkably similar to the relationship between *w* and the negative log-likelihood of our logistic regression of PNS (Figure 2b). We also estimated *α_mix_* values for each value of *w* (Figure 4c). Surprisingly, values of *w* that were lower than the best-fit value of *w*=0.96 produced larger absolute *α_mix_* estimates. This may be because at high values of *w*, *p_mix_* approaches 0 for some bins of SNPs, which results in overestimates of per-allele effect size variance under the *α_mix_* model, which cause best-fit estimates of *α_mix_* to be attenuated towards 0. This is analogous to the known phenomenon that the 𝛼 model overestimates per-allele effect size variance at low values of *p* according to evolutionary forward simulations, in which per-allele effect size variance plateaus at low values of *p* (Figure 2 of ref.^19^). To investigate further, we fit the *α_mix_* model while restricting to SNPs with *p_A_* ≥ 0.05 and *p_E_* ≥ 0.05. Accordingly, *α_mix_* estimates were less dependent on *w* when restricting to SNPs with *p_A_* ≥ 0.05 and *p_E_* ≥ 0.05 (best-fit *w*=0.94 and *α_mix_*=−0.22; Supplementπary Figure 10). Estimates of *α_mix_* were closer to 0, providing further evidence that SNPs with *p_A_* < 0.05 are the primary drivers of the European MAF-dependence of per-allele effect size variance.

**Figure 4:**
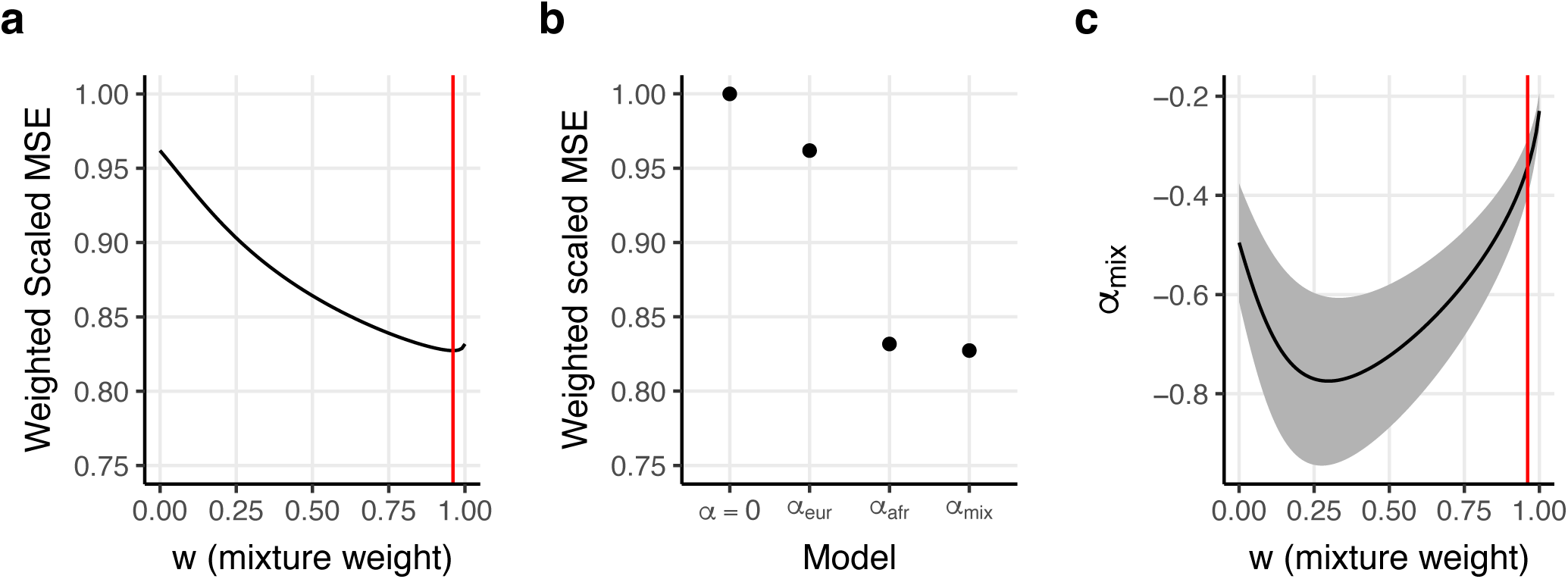
Results of fitting the *α_mix_* model across 50 diseases/traits. We report **(a)** the weighted scaled mean squared error with respect to *w*, **(b)** the weighted scaled mean squared error for various models, including a model without MAF dependence (𝛼=0), a model with European MAF dependence (𝛼_𝑒𝑢𝑟_), a model with African MAF dependence (𝛼_𝑎𝑓𝑟_), and the *α_mix_* model, and **(c)** estimates of *α_mix_* as a function of *w*. Weighted scaled MSE weights the MSE for each bivariate MAF bin by the number of SNPs it contains and scales the sum over all traits such that the null model has a weighted scaled MSE of 1. Red lines in (a) and (c) denote our point estimate of *w*. The shaded region in (c) denotes 95% confidence intervals for *α_mix_*. Numerical results are reported in Supplementary Table 10.

We estimated *α_mix_* and *w* parameters separately for each disease/trait. Results are reported in Figure 5, Supplementary Figures 11-12, and Supplementary Table 11. The random effects meta-analysis estimate of *w* was 1.00 (95% CI: (0.99, 1.00)) (similar to our joint estimate of 0.96; see above). Despite substantial noise in our estimates, *w* estimates were significantly greater than 0.5 for 24/50 traits (*P*<0.05/50; greater dependence on African MAF vs. European MAF) and significantly greater than 0 for 32/50 traits (*P*<0.05/50; significant dependence on African MAF)— but significantly less than 1 for 0/50 traits (*P*<0.05/50; significant dependence on European MAF). The random effects meta-analysis *α_mix_* estimate was =−0.31 (95% CI: (−0.26,−0.36)) (similar to our joint estimate of −0.34; see above). We determined that there was no statistically significant heterogeneity in *w* or *α_mix_* in a random effects meta-analysis (between-trait variance estimate of 0) or a fixed-effect meta-analysis (*P*=0.97 and 0.98 for *w* and *α_mix_*, respectively).

**Figure 5:**
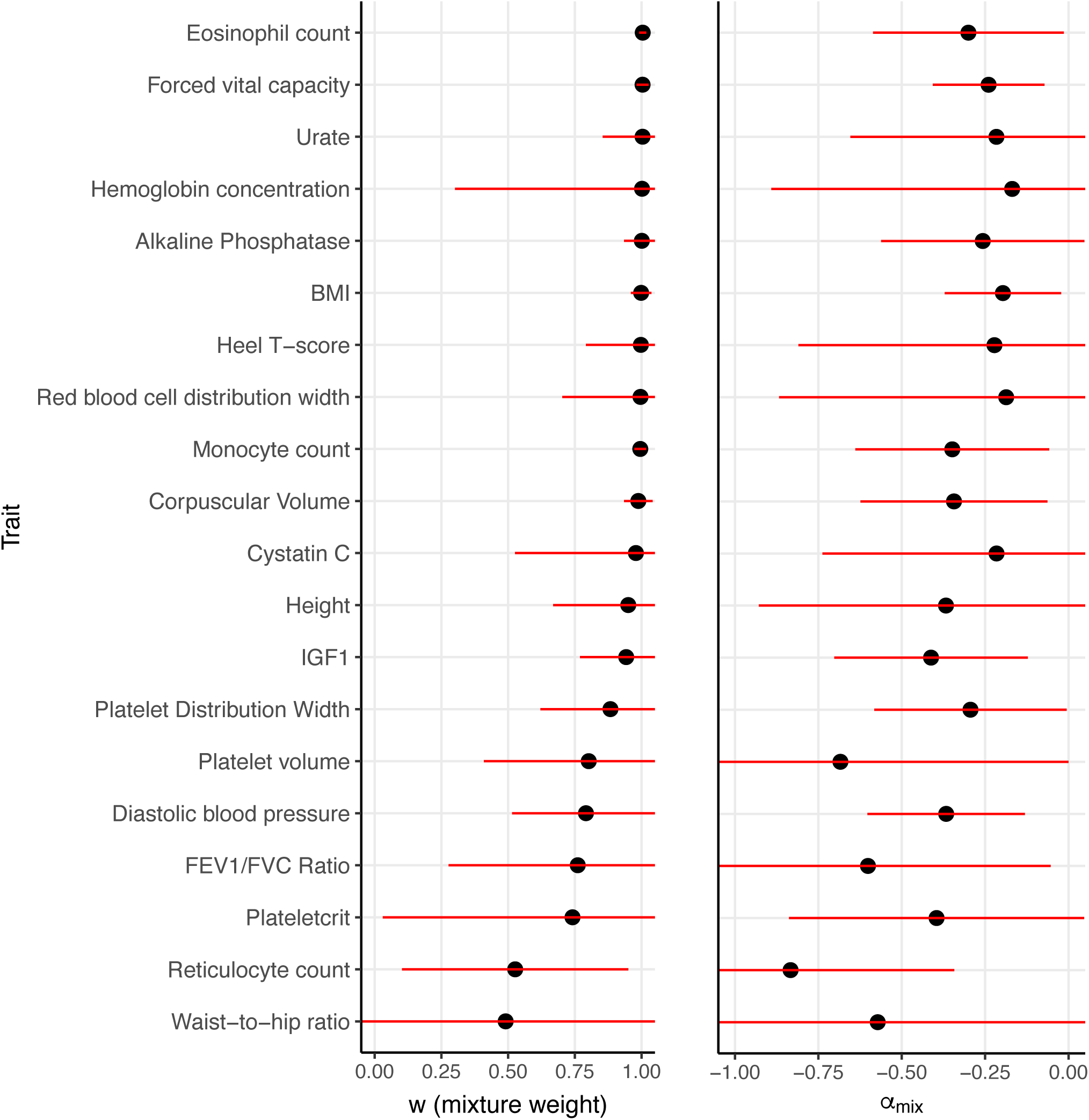
Results of fitting the *α_mix_* model separately for 20 out of 50 diseases/traits. We report estimates of **(a)** *w* and **(b)** *α_mix_*. Diseases/traits are ordered by point estimate of *w*. The 20 diseases/traits with highest *N*h^2^*point estimates are displayed. Supplementary Figure 11 displays results for all 50. Error bars denote 95% confidence intervals. Numerical results for all 50 traits are reported in Supplementary Table 11.

We performed 5 secondary analyses. First, we estimated *α_mix_* and *w* parameters separately for each disease/trait, while restricting to SNPs with *p_A_* ≥ 0.05 and *p_E_* ≥ 0.05 (Supplementary Figure 13). Our random effects meta-analysis estimates of *w*=0.53 (95% CI: (0.43, 0.63)) differed substantially from that of our main analysis, while our estimate of *α_mix_*=−0.33 (95% CI: (−0.19, −0.46)) was similar to the main analysis estimate. Identifying the reasons for this difference in *w* estimates when estimating parameters separately for each disease/trait and restricting to common SNPs is a direction for future research. Second, we fit the *α_mix_* model while estimating African MAF using a much smaller sample of Yoruba genomes (*N=*107 from 1000 Genomes YRI vs. *N*=46,779 African individuals and *N*=489 European individuals in the main analyses) (Supplementary Figure 14). Our estimates of *w*=0.96 (95% CI: (0.84, 1.07)) and *α_mix_*=−0.37 (95% CI: (−0.14, −0.59)) were very similar to those of the main analyses. Third, we fit the *α_mix_* model while estimating per-allele effect size variances using the standard baseline-LD (v2.2) model (Supplementary Figure 15), rather than our extension of the baseline-LD (v2.2) model which includes annotations for 120 bivariate MAF bins. Our estimates of *α_mix_*=−0.34 (95% CI: (−0.25, −0.44)) and *w*=0.85 (95% CI: (0.76, 0.95)) were similar to those of the main analysis. Fourth, to test whether African LD-related annotations in the baseline-LD (v2.2) model were falsely inducing African MAF dependence, we fit the *α_mix_* model while estimating per-allele effect size variances using 120 bivariate MAF bins rather than our extension of the baseline-LD (v2.2) model which includes all annotations from the standard baseline-LD (v2.2) model (Supplementary Figure 16). Our estimates of *w*=0.98 (95% CI: (0.86, 1.10)) and *α_mix_*=−0.38 (95% CI: (0, −0.76)) were very similar to those of the main analyses. Fifth, we fit the *α_mix_* model using two disjoint subsets of the 50 diseases/traits from the main analysis: 29 UK Biobank diseases/traits and 21 diseases/traits from publicly available summary statistics (Supplementary Figures 17-18). Results from each subset of diseases/traits were very similar to each other (e.g. *w*=0.97 (95% CI: (0.76, 1.17)) vs. *w*=0.96 (95% CI: (0.87, 1.05)), respectively).

### Simulations of *α_mix_* estimates

To test the validity of our *α_mix_* estimation procedure, we simulated quantitative GWAS traits according to the *α_mix_* model using real genotypes from 337,448 unrelated British individuals from UK Biobank and 5,907,305 SNPs with European MAF ≥ 0.05. We varied the true value of *w* between 0 and 1, set *α_mix_* to −0.38 (consistent with previous estimates in European populations^17–19^), set 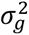 such that *h^2^*=0.5, set *p_mix_*=max(*w*\**p_A_* + (*1*-*w*)*p_E_, *T*) with *T*=0.005 (to avoid unrealistically large values of 𝐸[𝛽^2^|𝑝_𝐴_, 𝑝_𝐸_] for SNPs with low values of *p_A_*, consistent with ref.^19^; this implies that the generative model is different from the inference model), and set the number of causal SNPs to 10,000; other simulations settings were also evaluated. We fit the *α_mix_* model (Equation 1 and Methods) using a single *w* parameter and a single *α_mix_* parameter across groups of 25 traits (which resulted in total *N*h^2^*=4.2*10^6^, vs. 4.1*10^6^ in real trait analyses).

Results are reported in Figure 6 and Supplementary Table 12. We determined that our *w* estimates were conservative in the sense that they were biased away from extreme values, e.g. average estimate ranging from 0.16 when true *w*=0.00 to 0.97 when true *w*=1.00 (Figure 6a); estimates were approximately unbiased (0.94) at true *w*=0.95, which was closest to our estimate for real traits. Estimates of 𝛼 were biased downward (relative to true 𝛼=−0.38) at most values of *w*, e.g. −0.48 when *w*=0.00 and −0.42 when *w*=0.95 (but −0.35 when *w*=1.00) (Figure 6b). Secondary analyses provided below indicate that these biases are due to imperfect estimation of mean effect size variance via S-LDSC and misspecification of the inference model with respect to *T*.

**Figure 6:**
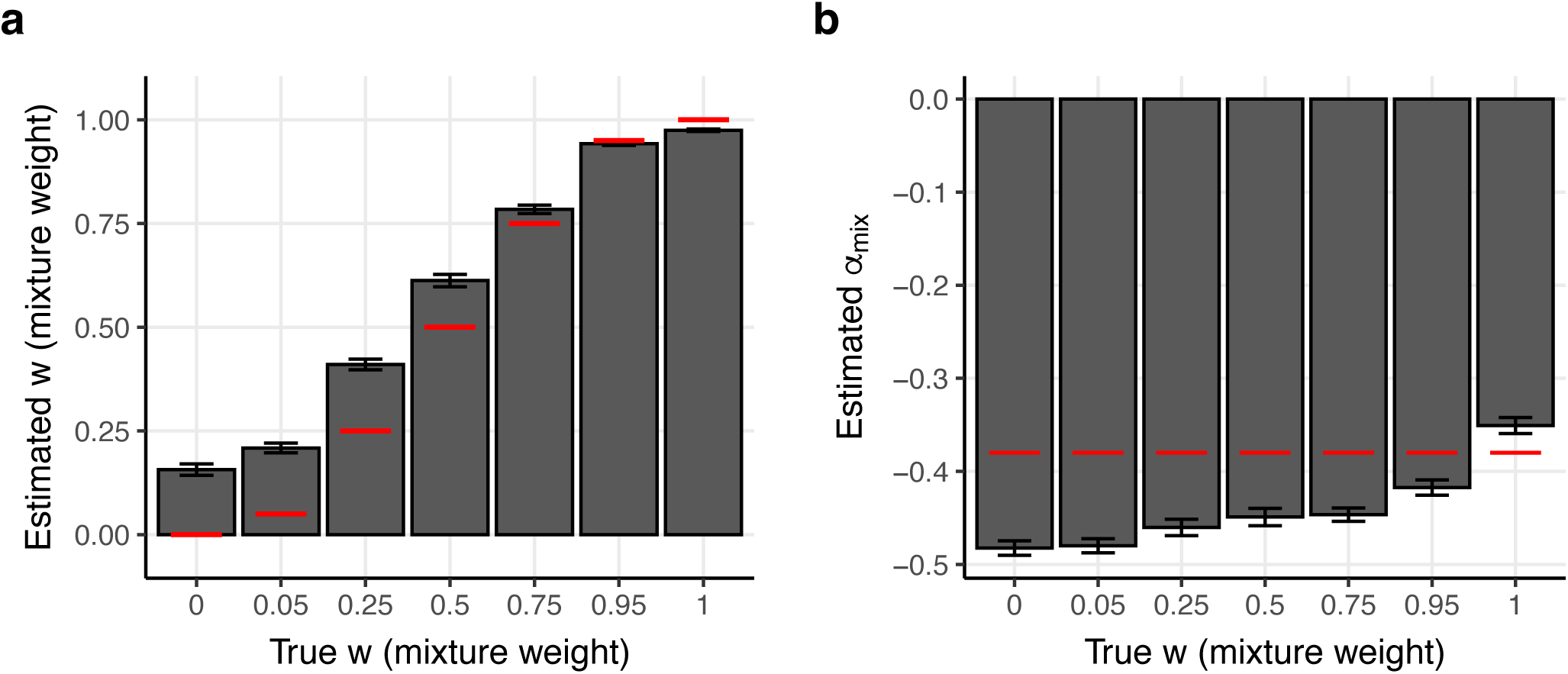
Results of fitting the *α_mix_* model in simulations. We report **(a)** estimates of *w* and **(b)** *α_mix_*, at different values of true *w*. Bars denote mean estimates across 40 simulation replicates (each analyzing 25 traits). Red lines denote true parameter values. Error bars denote 95% confidence intervals. Numerical results are reported in Supplementary Table 12.

We performed 6 secondary analyses. First, we simulated effects based on MAF from *N*=337,448 unrelated British individuals, but performed inference using MAF from *N*=489 European individuals from 1000 Genomes to simulate the effect using noisy MAF estimates (Supplementary Figure 19). Results were very similar to those of the main analyses. Second, we varied *T*, the MAF threshold used by the generative model (Supplementary Figures 20-21), which determines how high 𝐸[𝛽^2^|𝑝_𝐴_, 𝑝_𝐸_] is for SNPs with low values of *p_A_*. Results with *T*=0.05 were similar to those of the main analyses, but with upward bias (rather than downward bias) in our *α_mix_* estimates at *w*=0.75 and *w*=0.95 and slightly greater upward bias in *α_mix_* at *w*=1.00. Results with *T*=1.5*10^-5^ (equivalent to a minor allele count of 1) were similar to those of the main analyses, but with less bias in our estimates of *w* at high values of *w*, and downward bias (rather than upward bias) in our estimates of *α_mix_* at *w*=1.00. We conclude that our estimates of *w* are insensitive to plateauing of E[𝛽^2^|𝑝_𝐴_, 𝑝_𝐸_] at low values of *p_A_*. Third, we varied the number of causal SNPs per trait (Supplementary Figure 22-23). With 5,000 (instead of 10,000) causal variants per trait, results were similar to the main analyses, with slight downward bias at *w*=1. With 20,000 causal variants per trait, results were very similar to the main analyses. Fourth, we applied our inference procedure while using true values of 𝛽^2^ (instead of estimates of mean effect size variance from S-LDSC; Supplementary Figure 24). Estimates of *w* and *α_mix_* were approximately unbiased, except for at *w* ≥ 0.95, where estimates of *α_mix_* were biased upwards. We conclude that in our main simulations, bias in estimates of *w* and bias in estimates of *α_mix_* at *w* ≤ 0.75 were caused by our approximation of mean effect size variance. Fifth, we applied our inference procedure while using the true values of 𝛽^2^ (instead of estimates of mean effect size variance from S-LDSC), restricting the inference procedure to causal variants, and thresholding *p_mix_* for individual SNPs at the true value of *T* (Supplementary Figure 25). We also performed inference using the same approach, but without thresholding *p*_mix_ (Supplementary Figure 26). Estimates of *α_mix_* were biased upwards at *w* ≥ 0.95 when *p_mix_* was not thresholded, and were unbiased when *p_mix_* was thresholded at *T*. We conclude that the bias in our estimates of *α_mix_* at *w* ≥ 0.95 in the main simulations was due to model misspecification with respect to *T*. Finally, we estimated trait-specific values of *w* and *α_mix_*, rather fitting a joint model across all 25 traits (Supplementary Figure 27). Results were very similar to the main analyses, but with greater uncertainty in mean parameter estimates.

## Discussion

We have shown that per-allele disease and complex trait effect sizes of SNPs that are common in European populations are predominantly a function of African MAF, not European MAF. This represents a significant departure from standard approaches for modeling MAF dependence, where per-allele effect sizes estimated in a given population are related to MAF in the same population^4,9,15–27,29–33,35–38^. We have provided two lines of evidence in favor of an African MAF-dependence model. First, we showed that non-synonymous variant status is much better predicted using African MAF than European MAF. This approach requires very few assumptions and is likely robust to model misspecification, but is limited in that it only focuses a single, relatively narrow subset of human genetic variation. Second, we used disease/trait GWAS summary statistic data from European populations to show that per-allele effect size variance is also much better predicted using African MAF than European MAF. This approach reached very similar conclusions to our analysis of non-synonymous variation.

Our work is motivated in part by three recent population genetic studies exploring the relationship between African MAF, European MAF, and GWAS effect sizes. Refs. ^28,34^ performed simulations exploring the effect of bottlenecks on genetic architectures and MAF dependence, using models in which per-allele effect sizes in one population are dependent on MAF in another (ref. ^28^ focuses specifically on the out-of-Africa bottleneck, while ref. ^34^ does not). However, neither study analyzed empirical GWAS data or demonstrated that this model fits empirical data. Ref. ^39^ used conditional allele frequency spectra to differentiate modes of selection on GWAS-associated variants (emphasizing joint analyses of African MAF and European MAF, as we do in this study), but did not quantify the dependence of per-allele effect sizes on African MAF or European MAF.

Our work has important implications both for analyses of European GWAS data and, more generally, populations of all ancestries. Modeling MAF-dependent architectures has been shown to impact studies of genetic architectures and negative selection^2,4,5,17–24,26,28,29,31,33,34,37–39,49–51,53,59,60^, polygenic risk prediction^9,11,61^, and fine-mapping^16,61^, and our results suggest that specifically modeling African MAF-dependent architectures will improve all of these endeavors, not only in European populations (the focus of this study) but in population of all ancestries as more GWAS data in diverse populations becomes available^62–67,42,68^.

Our work has several limitations. First, our analyses focus exclusively on African and European MAF. We expect that drift from the out-of-Africa bottleneck causes MAF in all non-African ancestries to be less informative for per-allele effect size variance^44^, although we have not explicitly tested this hypothesis. Second, our analyses of diseases/traits focus exclusively on European-ancestry GWAS data, which limits our disease/trait analyses to SNPs with *p_E_* > 0.05; however, our analysis of non-synonymous variant status spans a larger set of SNPs (*p_A_* > 0.002 and/or *p_E_* > 0.002), providing some evidence that our findings are generalizable. Analyses of diseases/traits in non-European populations will be more informative as more GWAS data in diverse populations becomes available^62–67,42,68^. Third, our analyses focus on continental ancestries and do not consider subcontinental ancestries with distinct allele frequencies, demographic histories, and selective forces^45–47,69,70^; however, allele frequency differences between populations of the same continental ancestry are generally smaller than allele frequency differences between populations of different continental ancestry^71^. Fourth, our estimate of the degree of European MAF-dependence under the 𝛼 model instead of the *α_mix_* model (𝛼=−0.50, 95% CI: (−0.61,−0.38), Figure 4c) was stronger than refs.^17–19^ (𝛼 = −0.38 to −0.36 meta-analysis across traits) (though weaker than refs.^20,33^; 𝛼 = −0.65 to −0.58). When estimating 𝛼 using S-LDSC with the standard Baseline-LD (v2.2) model, which does not include African MAF annotations, (Supplementary Figure 15), we obtain an estimate that is consistent with refs.^17–19^ (𝛼 = −0.41, 95% CI (−0.51, −0.32)); regardless of the S-LDSC model used, we find that per-allele effect size variances are predominantly African MAF-dependent, which is our main conclusion. Fifth, our approach to fitting the *α_mix_* model suffers some biases in simulations (Figure 6); however, estimates of *w* were conservative (in the sense that they were biased away from extreme values), and approximately unbiased for values of *w* close to our estimate of *w*=0.96 for real traits, thus providing strong support for our conclusion that per-allele effect sizes are predominantly African MAF-dependent. Sixth, we did not perform simulations to assess whether the *α_mix_* model is consistent with evolutionary models at appropriate parameter settings; a previous study performed forward simulations^72^ to show that the 𝛼 model is generally consistent with an evolutionary model involving a single population^19^ (except that per-allele effect size variance plateaus at low values of minor allele frequency; Figure 2 of ref.^19^), and extending those forward simulations to incorporate African and non-African populations (impacted by the out-of-Africa bottleneck) is an important future research direction. Seventh, analogous to previous studies using the 𝛼 model^4,18,19,21–26,29,31,32,34,35,38^, our analyses using the *α_mix_* model do not account for non-infinitesimal disease/trait effects^73^; we note that failure to model non-infinitesimal effects when estimating SNP-heritability leads to suboptimal precision, but does not induce bias^73^. Eighth, African MAF is unlikely to be informative of per-allele effects among SNPs which are younger than the out-of-Africa bottleneck and which are common in European ancestry populations, although these represent a small fraction of European common variation; modeling MAF-dependence as a function of allele age is a potentially impactful future research direction. Despite these limitations, we have shown that per-allele disease and complex trait effect size variances are predominantly African MAF-dependent in European populations.

## Data Availability

All data produced in the present study are available upon reasonable request to the authors.

## Supplementary Tables

**Supplementary Table 1: Overview of 50 diseases/complex traits analyzed.** We report the SNP heritability (h^2^) as estimated by LDSC and GWAS sample size.

(see .xlsx file)

**Supplementary Table 2: The proportion of non-synonymous SNPs in a grid of bivariate African and European MAF bins.** We report the proportion of non-synonymous SNPs, the proportion of synonymous SNPs, the number of SNPs, and the Wald standard error on the proportion of non-synonymous SNPs.

(see .xlsx file)

**Supplementary Table 3: The proportion of non-synonymous SNPs in African MAF deciles stratified by European MAF bins.** We report, for each decile, the African MAF, the proportion of non-synonymous variants, 95% confidence intervals on the proportion of non-synonymous variants, and the European MAF bin. We restricted to variants with *p_A_* ≥ 0.002

(see .xlsx file)

**Supplementary Table 4: Predictions from logistic regressions of SNP non-synonymous status on African MAF stratified by European MAF bins.** We report the predicted proportion of non-synonymous variants (and 95% confidence intervals) for a grid of 30,000 African MAF values from 0.002 - 0.5. In the regression African MAF was log transformed. We restricted to variants with *p_A_* ≥ 0.002.

(see .xlsx file)

**Supplementary Table 5: The proportion of non-synonymous SNPs in European MAF deciles stratified by African MAF bins.** We report, for each decile, the European MAF, the proportion of non-synonymous variants, 95% confidence intervals on the proportion of non-synonymous variants, and the African MAF bin. We restricted to variants with *p_E_* ≥ 0.002

(see .xlsx file)

**Supplementary Table 6: Predictions from logistic regressions of SNP non-synonymous status on European MAF stratified by African MAF bins.** We report the predicted proportion of non-synonymous variants and 95% confidence intervals for a grid of 30,000 European MAF values from 0.002 - 0.5. In the regression European MAF was log transformed. We restricted to variants with *p_E_* ≥ 0.002.

(see .xlsx file)

**Supplementary Table 7: Best-fit parameters for a logistic regression of non-synonymous status on log(*p_mix_*).** We report the log-likelihood, 𝛾 estimate, 𝛾 standard error, and 95% confidence interval on 𝛾 across 101 values of *w* from 0 to 1.

(see .xlsx file)

**Supplementary Table 8: Per-allele effect size variance across 50 diseases/traits for a grid of bivariate African and European MAF bins.** We report point estimates and standard errors of the meta-analyzed scaled mean effect size variance for each bin.

(see .xlsx file)

**Supplementary Table 9: Per-allele effect size variance across 50 diseases/traits as a function of MAF quintiles, stratified by African MAF.** We report, for each MAF quintile, the mean MAF, point estimate of the meta-analyzed scaled mean effect size variance, standard error of the meta-analyzed scaled mean effect size variance, and 95% confidence interval on the meta-analyzed scaled mean effect size variance.

(see .xlsx file)

**Supplementary Table 10: Results of fitting the *α_mix_* model across 50 diseases/traits.** We report point estimates and standard errors on *α_mix_* and point estimates and standard errors of the loss function across 101 values of *w* from 0 to 1.

(see .xlsx file)

**Supplementary Table 11: Results of fitting the *α_mix_* model separately for 50 diseases/traits.** We report point estimates and standard errors on *w*, *α_mix_*, and the loss function for each of 50 diseases/traits.

(see .xlsx file)

**Supplementary Table 12: Results of fitting the *α_mix_* model in simulations.** We report point estimates and standard errors on the mean estimate of *w* and *α_mix_*.

(see .xlsx file)

## Supplementary Figures

**Supplementary Figure 1:**
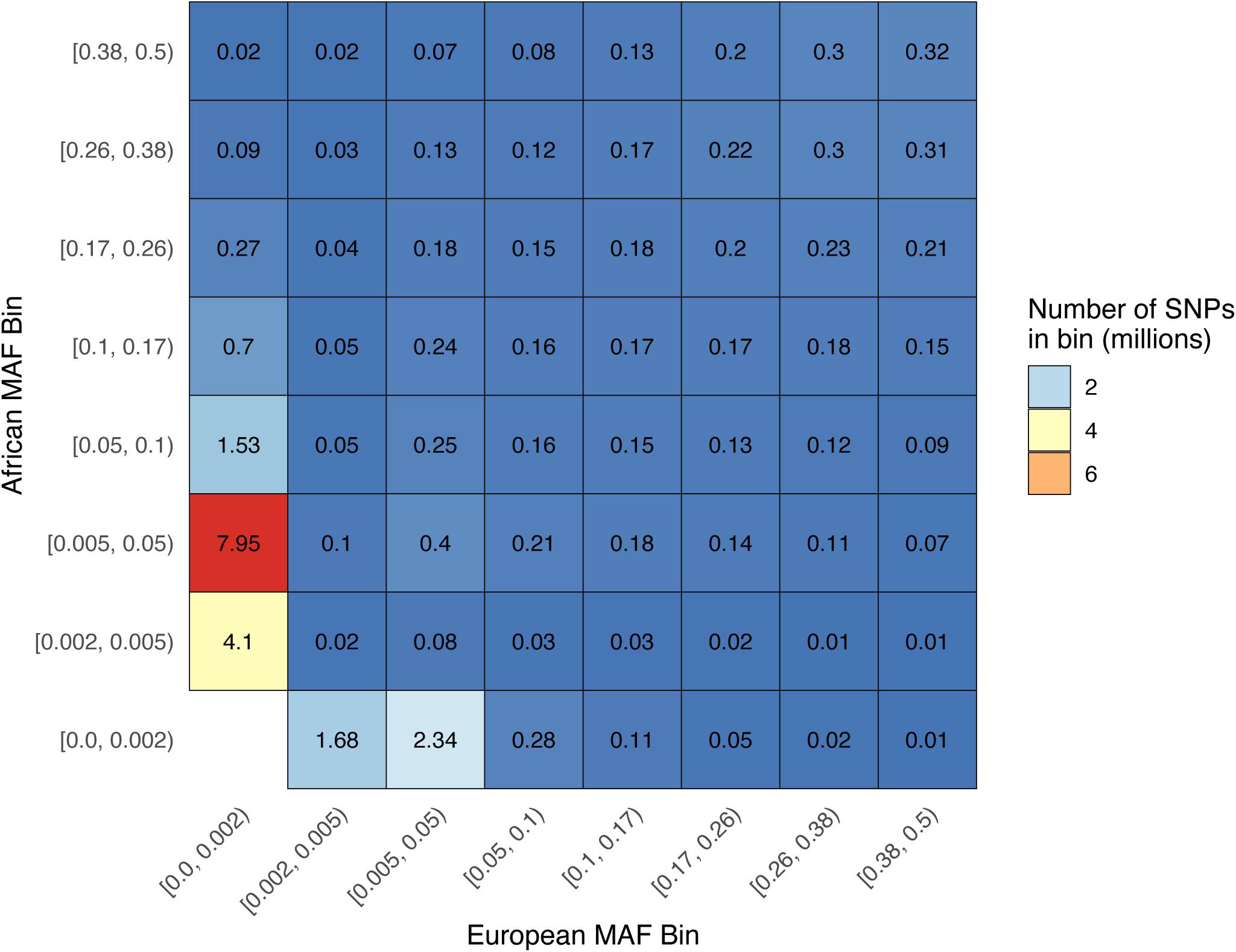
Number of SNPs in each AoU MAF stratum. We report the number of SNPs contained in each grid element given in Figure 1a. Each grid element contains SNPs with unadmixed African and European MAF that fall in a pair of MAF intervals. A justification for MAF interval boundaries is given in Methods.

**Supplementary Figure 2:**
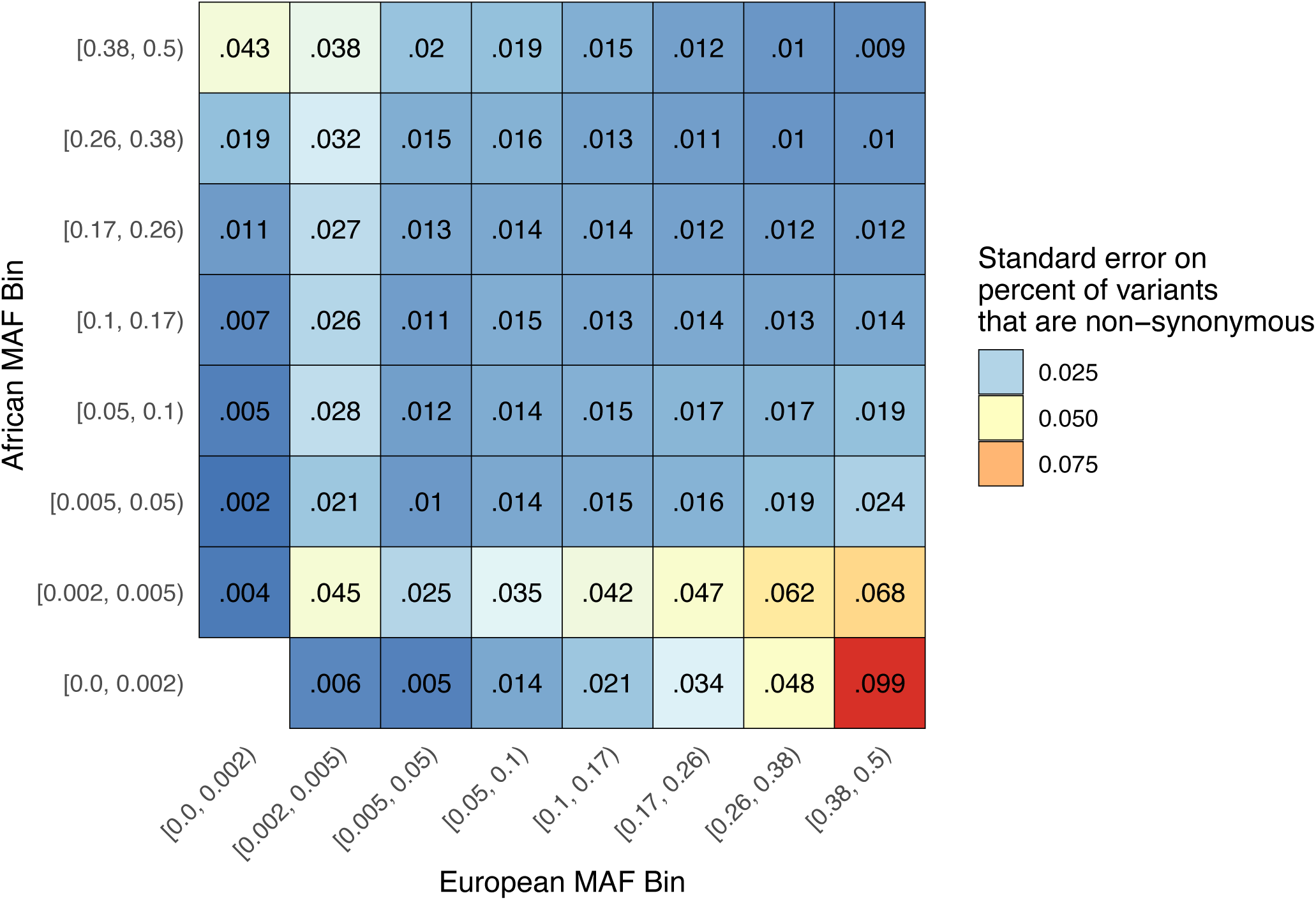
Standard error on the proportion of non-synonymous SNPs in each MAF stratum. We report the binomial proportion standard error for each grid element given in Figure 1a. Each grid element contains SNPs with unadmixed African and European MAF that fall in a pair of MAF intervals. A justification for MAF interval boundaries is given in Methods.

**Supplementary Figure 3:**
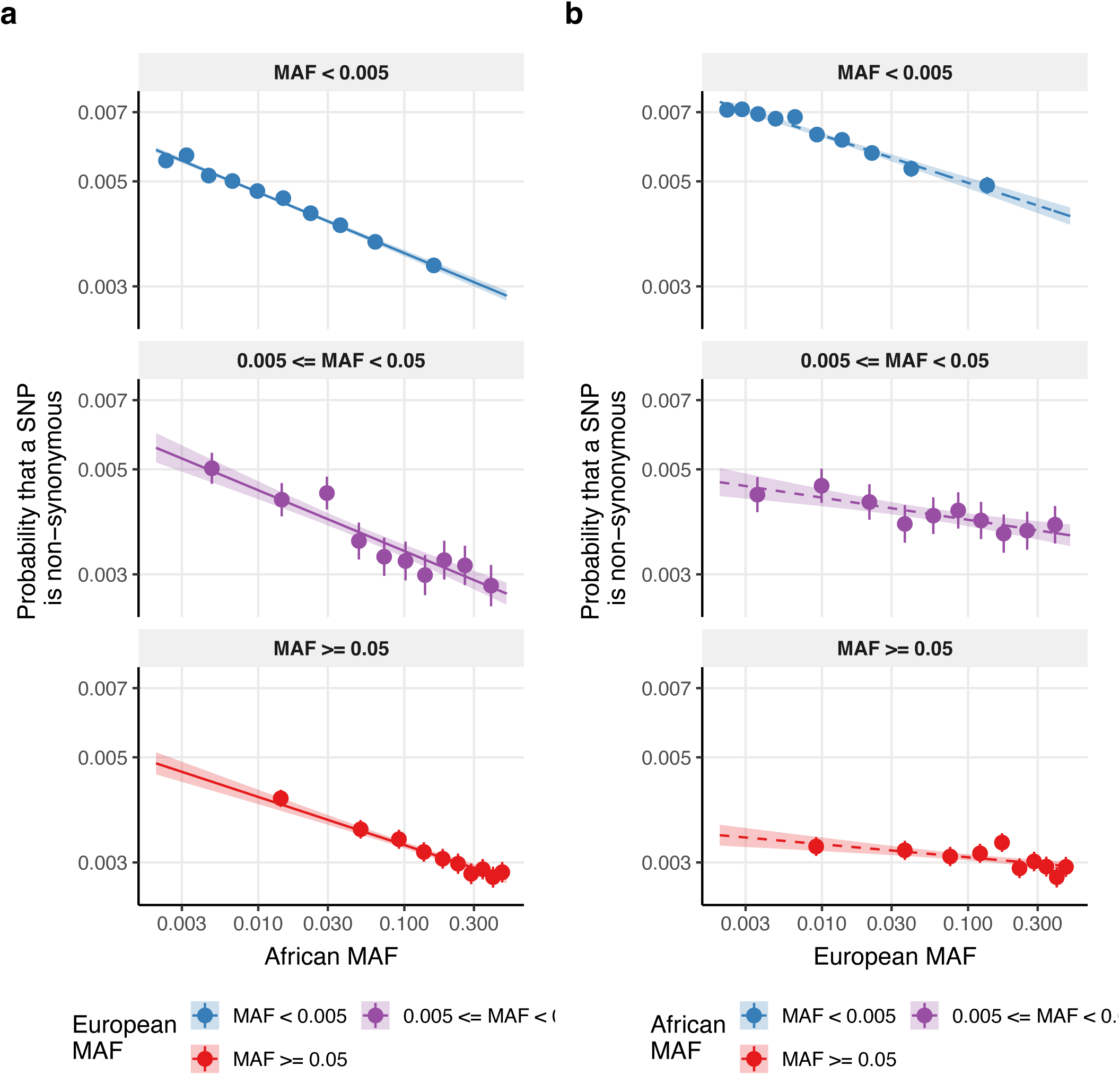
Logistic regression for non-synonymous status as a function of unadmixed African and European minor allele frequencies separated by MAF category. We report **(a)** the probability that a SNP is non-synonymous as a function of African MAF after stratifying by European MAF categories (for SNPs with *p_A_* ≥ 0.002) as predicted by logistic regression, and **(b)** the probability that a SNP is non-synonymous as a function of European MAF after stratifying by African MAF categories (for SNPs with *p_E_* ≥ 0.002) as predicted by logistic regression. Error bars denote 95% Agresti-Coull confidence intervals. Shaded regions in (b) and (c) denote 95% confidence intervals on the mean estimate. Logistic regression fits match non-parametric MAF deciles remarkably well, indicating that our conclusions are unlikely to be impacted by model misspecification.

**Supplementary Figure 4:**
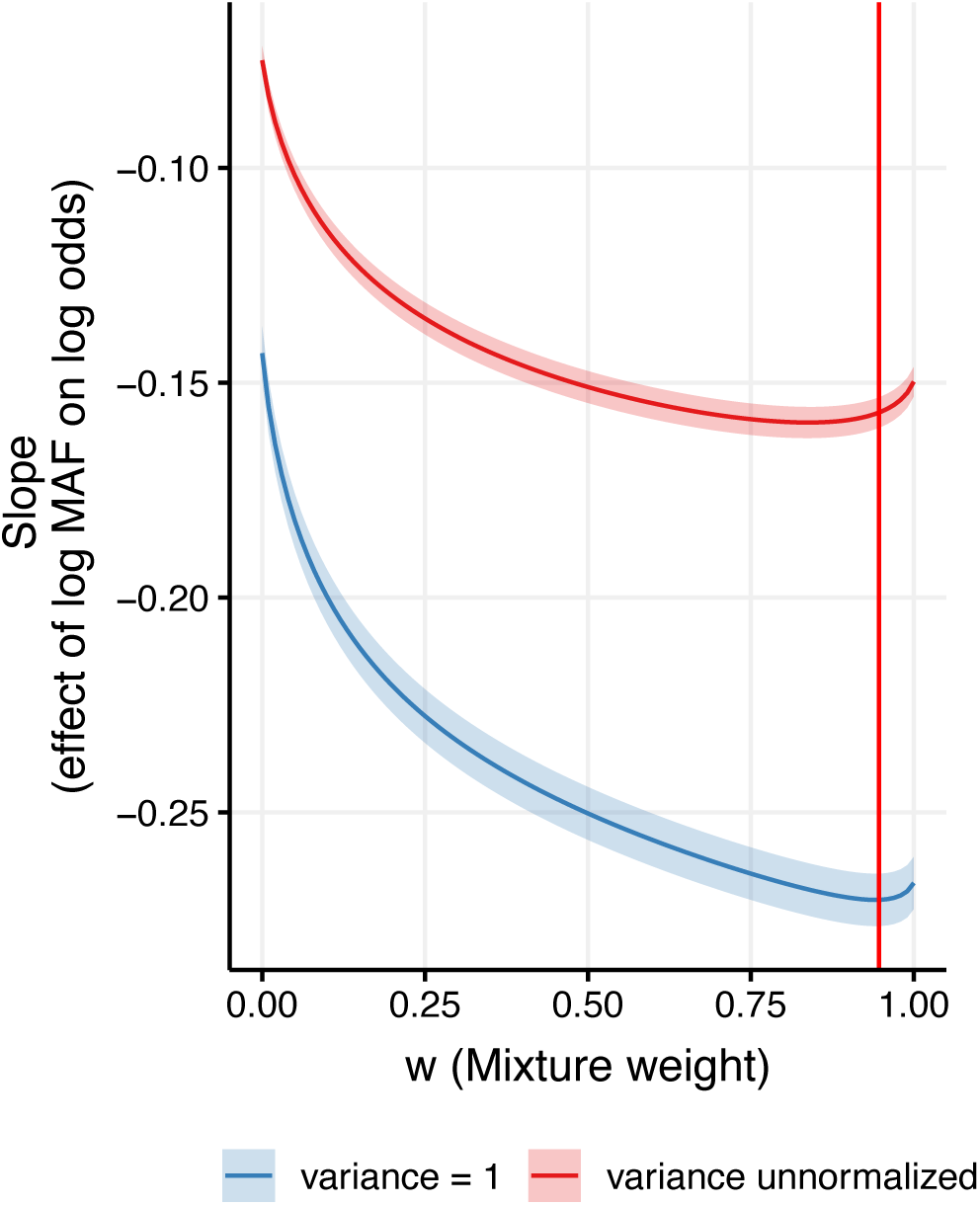
Best fit parameter for a logistic regression of non-synonymous status on log(*p_mix_*) while scaling log(*p_mix_*) to have unit variance. We report the regression slope (𝛾) for values of w between 0 and 1. The blue curve denote results for a logistic regression on log(*p_mix_*) while scaling log(*p_mix_*). The red curve denotes results for a logistic regression on log(*p_mix_*) without scaling log(*p_mix_*). The vertical red line denotes the MLE of *w=*0.95 (the MLE of *w* is not affected by scaling log(*p_mix_*)). With scaling, *w=*0.94 maximizes the absolute slope. Without scaling, *w*=0.84 maximizes the absolute slope.

**Supplementary Figure 5:**
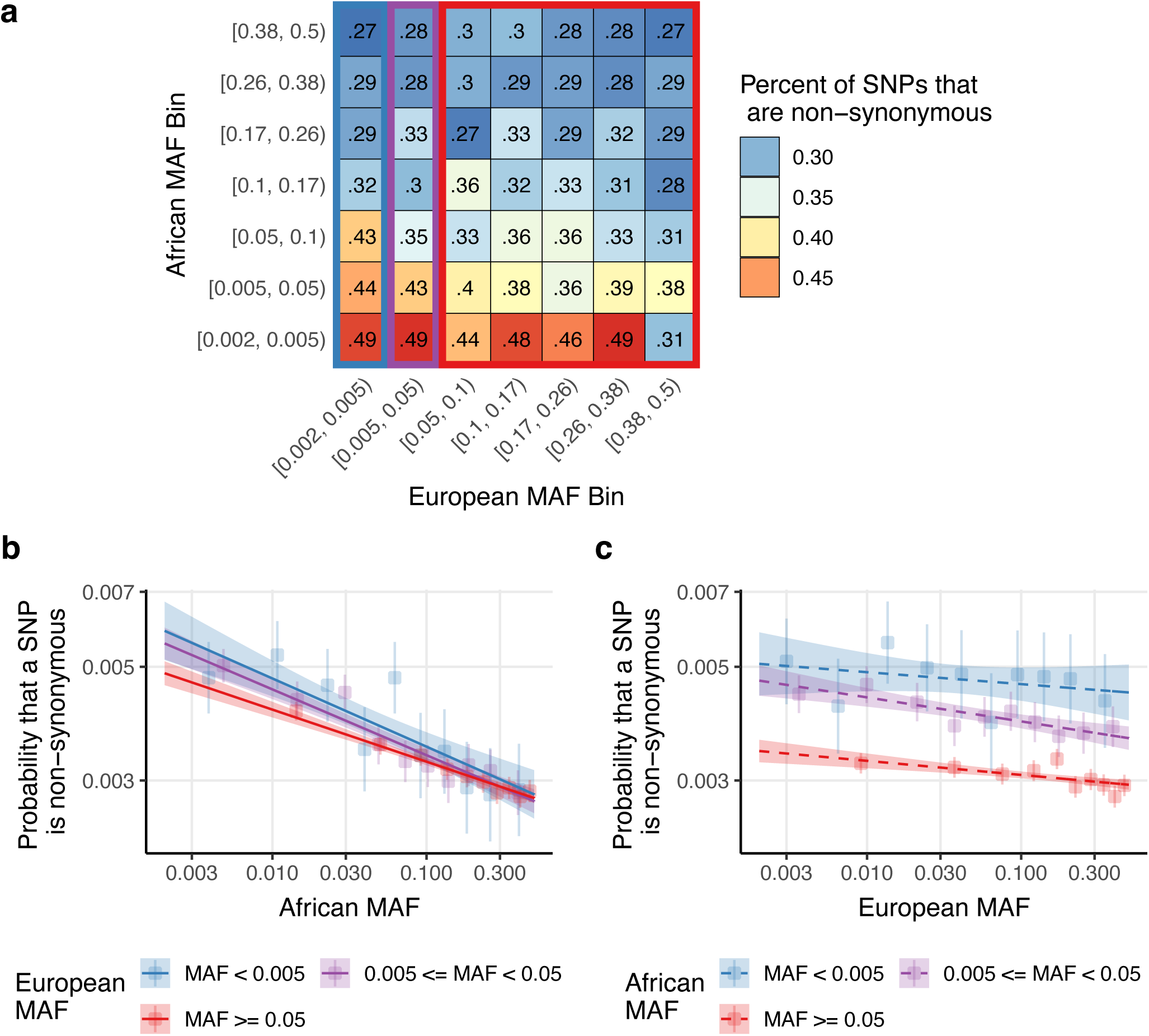
The proportion of non-synonymous SNPs as function of unadmixed African and European minor allele frequencies, restricted to variants with *p_A_* ≥ 0.002 and *p_E_* ≥ 0.002: We report **(a)** the percentage of SNPs that are non-synonymous in bivariate bins defined by unadmixed African MAF (rows) and European MAF (columns), **(b)** the probability that a SNP is non-synonymous as a function of African MAF after stratifying by European MAF categories (for SNPs with *p_A_* ≥ 0.002 and *p_E_* ≥ 0.002) as predicted by logistic regression, and **(c)** the probability that a SNP is non-synonymous as a function of European MAF after stratifying by African MAF categories (for SNPs with *p_E_* ≥ 0.002 and *p_A_* ≥ 0.002) as predicted by logistic regression. The blue, purple, and red rectangles in (a) correspond to the SNPs included in the blue, purple, and red lines in (b). The choice of MAF bin boundaries is discussed in Methods. Shaded regions in (b) and (c) denote 95% confidence intervals on the mean estimate. Error bars denote 95% Agresti-Coull confidence intervals.

**Supplementary Figure 6:**
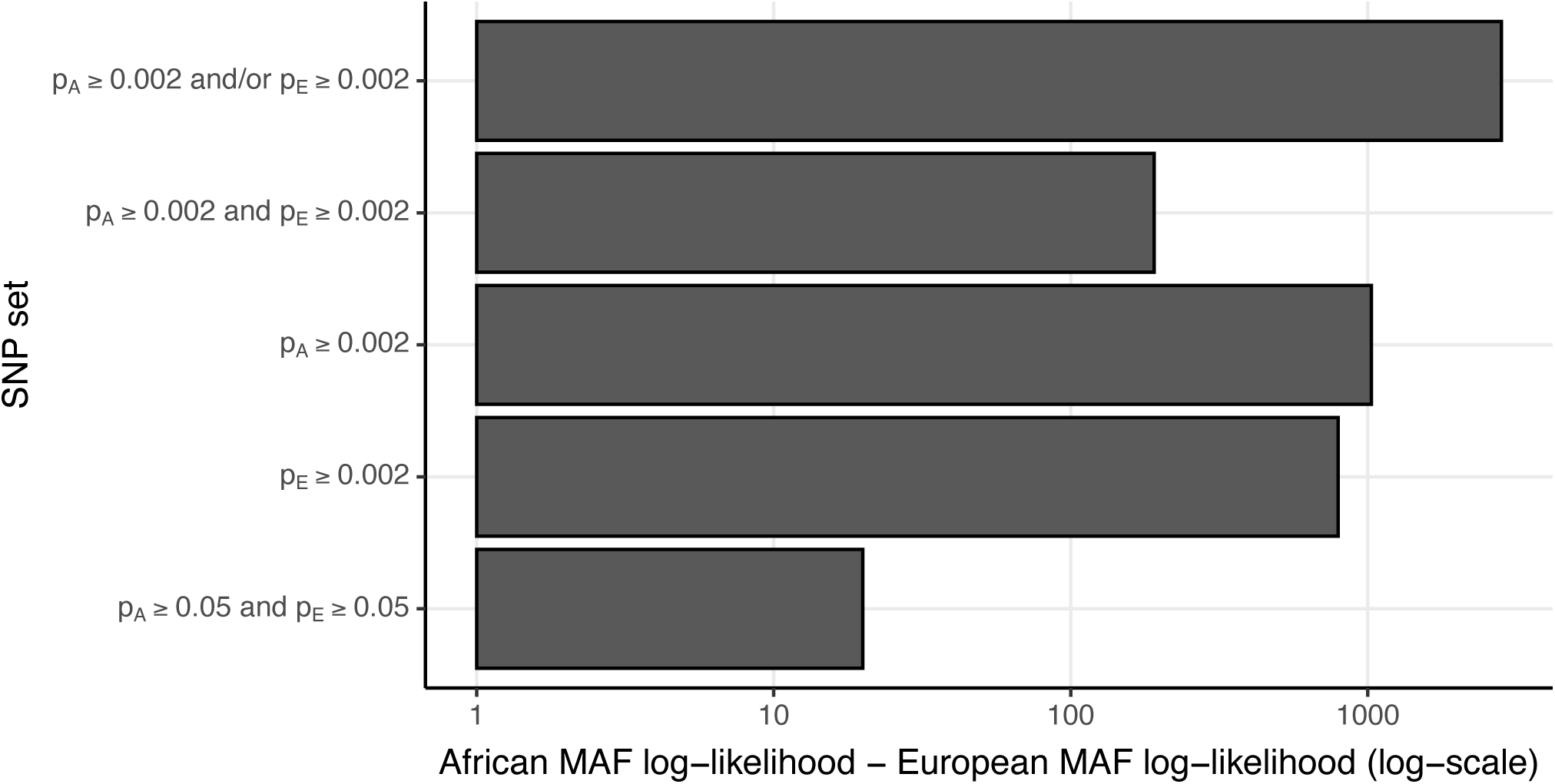
Difference in log-likelihood for a logistic regression of non-synonymous status on African MAF vs. a logistic regression of non-synonymous status on European MAF. We report the log-likelihood for a logistic regression of PNS on log(*p_A_*) minus the log-likelihood for a logistic regression of PNS on log(*p_E_*), while varying the SNP set. All log-likelihood differences were significant via Vuong’s test. For each SNP set, the exact same set of SNPs were included in the African MAF-only regression and the European MAF-only regression. For all SNPs, we thresholded MAF values at 0.002 after filtering for SNP set. That is, if *p_A_* < 0.002, *p_A_* was set to 0.002, and if *p_E_* < 0.002, *p_E_* was set to 0.002 (after restricting to the set of SNPs to be included in each SNP set).

**Supplementary Figure 7:**
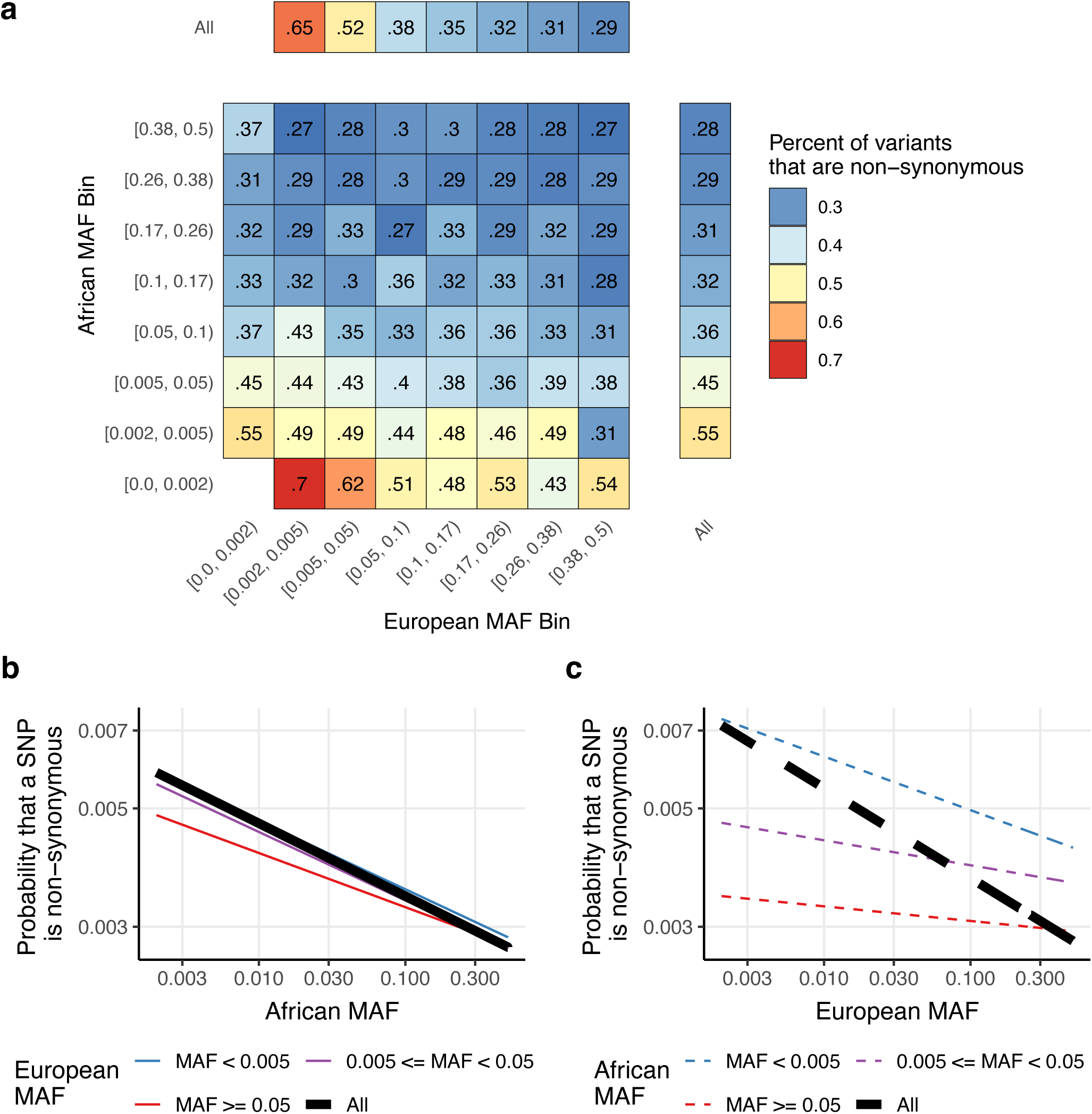
The proportion of non-synonymous SNPs as function of unadmixed African and European minor allele frequencies, highlighting the marginal effects of African MAF for SNPs with *p_A_* ≥ 0.002 and European SNPs with *p_E_* ≥ 0.002: We report **(a)** the percentage of SNPs that are non-synonymous in univariate bins defined by unadmixed African MAF (rightmost column) and European MAF (top row) and bivariate bins defined by unadmixed African MAF (other rows) and European MAF (other columns), **(b)** the probability that a SNP is non-synonymous as a function of African MAF marginally (black) and after stratifying by European MAF categories (colored lines) (for SNPs with *p_A_* ≥ 0.002) as predicted by logistic regression, and **(c)** the probability that a SNP is non-synonymous as a function of European MAF marginally (black) and after stratifying by African MAF categories (colored lines) (for SNPs with *p_E_* ≥ 0.002) as predicted by logistic regression.

**Supplementary Figure 8:**
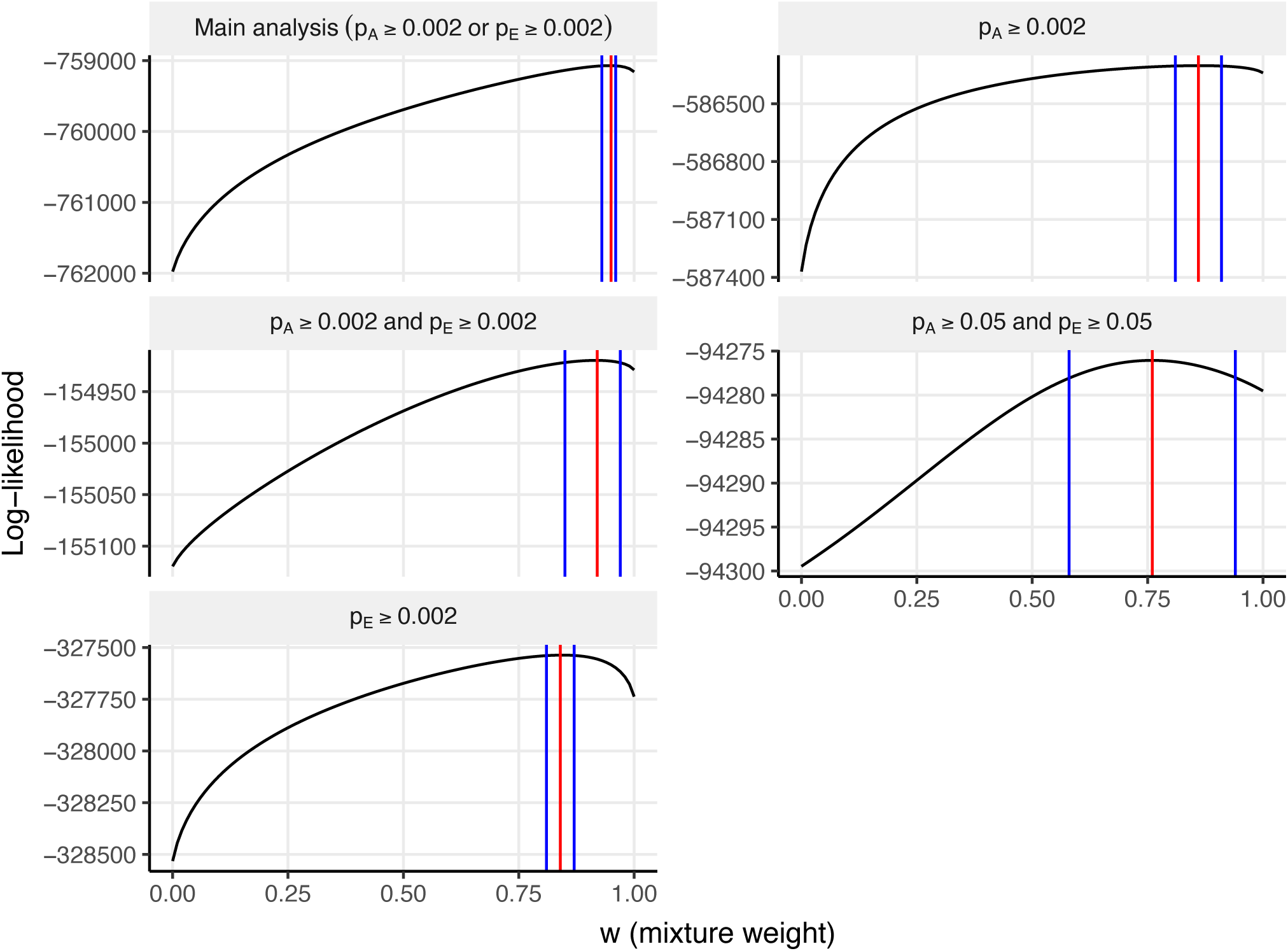
Log-likelihood for a logistic regression of non-synonymous status on log(*p_mix_*), varying the set of SNPs analyzed. We report the profile log-likelihood with respect to *w*, treating the regression intercept and slope as nuisance parameters. Each curve corresponds to a distinct set of SNPs included while fitting the regression. Red lines denotes the MLE for *w*. Blue lines denote 95% confidence interval bounds for *w*.

**Supplementary Figure 9:**
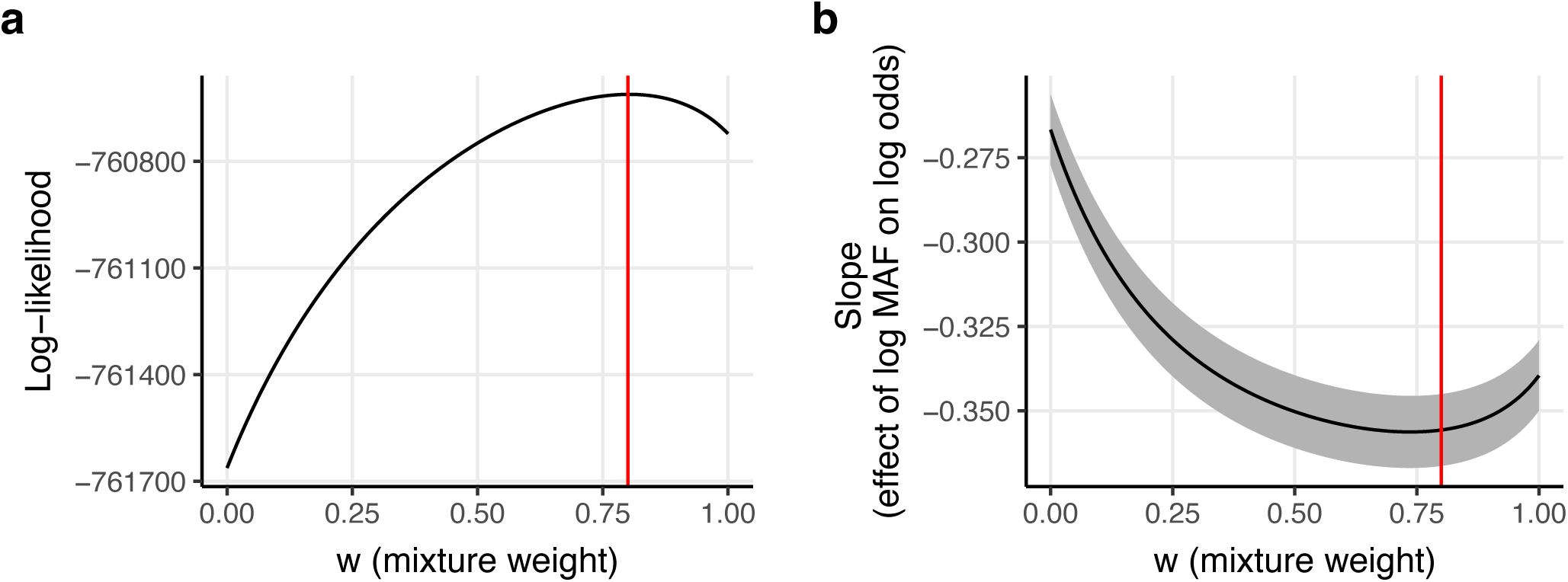
Best-fit parameters for a logistic regression of non-synonymous status on log(*p_mix_*), thresholding MAF at an alternate value of 0.05. We report **(a)** the profile log-likelihood with respect to *w*, treating the regression intercept and slope as nuisance parameters, thresholding *p_A_* and *p_E_* at 0.05, and **(b)** the regression slope (𝛾) for values of *w* between 0 and 1, thresholding *p_A_* and *p_E_* at 0.05. If we “threshold” a SNP at a MAF of *c*, it means that if a SNP has *p_A_* < c, we set *p_A_* to c, and if a SNP has *p_E_* < c, we set *p_E_* to c. In the main analyses, we threshold all MAF values at 0.002 (after filtering). Red lines in (a) and (b) denote the MLE estimate of *w*. The shaded region in (b) denotes 95% confidence intervals for 𝛾.

**Supplementary Figure 10:**
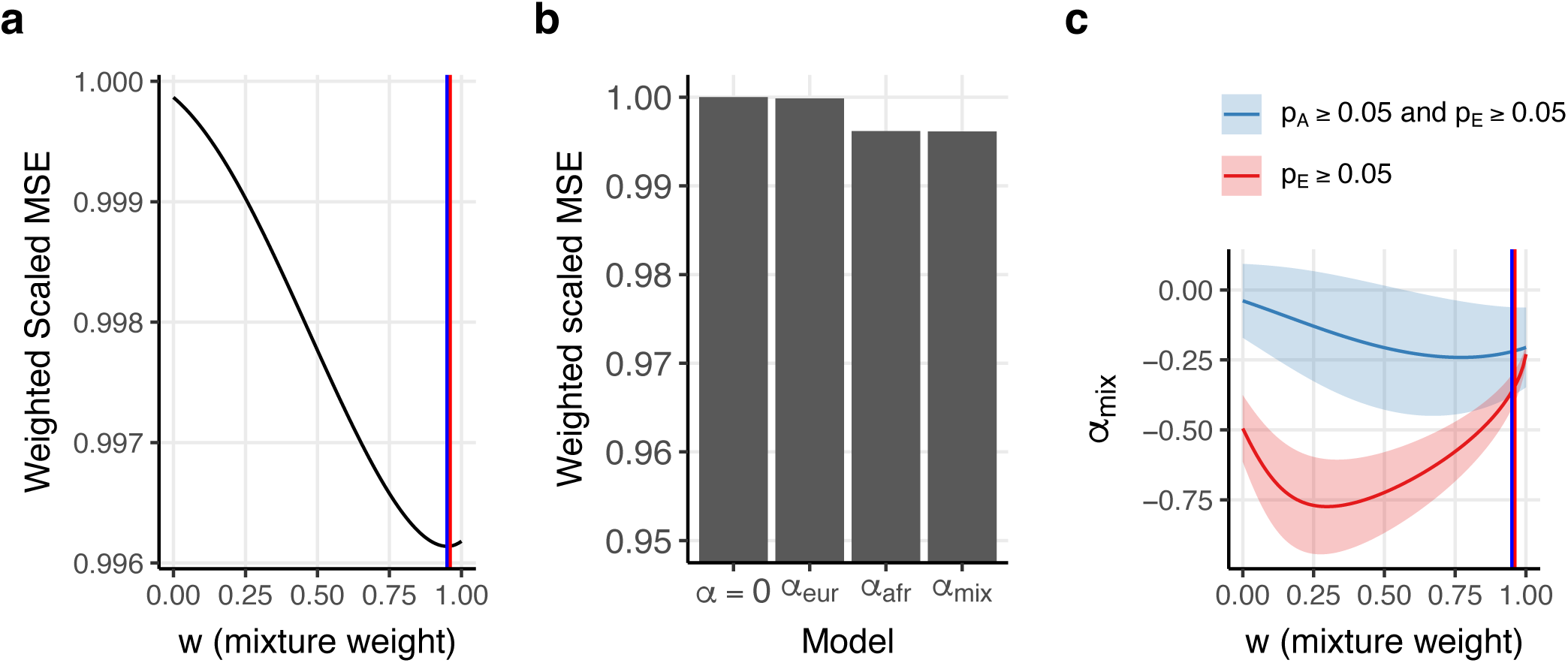
Results of fitting the *α_mix_* model across 50 diseases/traits restricted to variants with *p_A_* ≥ 0.05 and *p_E_* ≥ 0.05. We report **(a)** the weighted scaled mean squared error with respect to *w*, **(b)** the weighted scaled mean squared error for various models, including a model without MAF dependence (𝛼=0), a model with European MAF dependence (𝛼_𝑒𝑢𝑟_), a model with African MAF dependence (𝛼_𝑎𝑓𝑟_), and the *α_mix_* model, and **(c)** estimates of *α_mix_* as a function of *w*. Weighted scaled MSE weights the MSE for each bivariate MAF bin by the number of SNPs it contains and scales the sum over all traits such that the null model has a weighted scaled MSE of 1. Vertical lines in (a) and (c) denote point estimates of *w*. Blue lines correspond to estimates restricted to SNPs with *p_A_* ≥ 0.05 and *p_E_* ≥ 0.05 and red lines correspond to estimates from the main analyses (restricted to SNPs with *p_A_* ≥ 0.05). Shaded regions in (c) denote 95% confidence intervals.

**Supplementary Figure 11:**
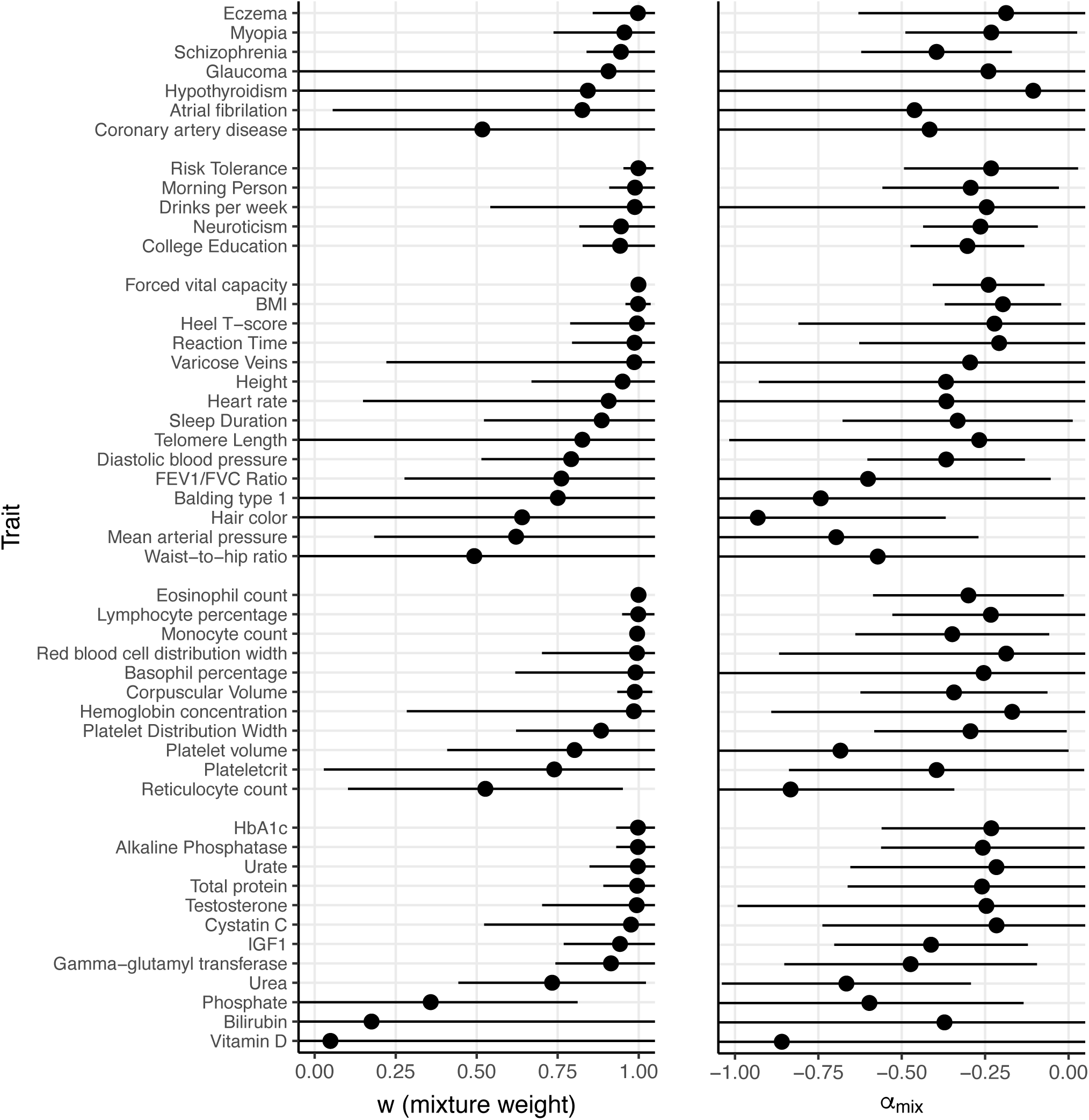
Results of fitting the *α_mix_* model separately for 50 diseases/traits. We report estimates of **(a)** *w* and **(b)** *α_mix_*. Diseases/traits are ordered by point estimate of *w* within disease/trait categories. Error bars denote 95% confidence intervals. Numerical results for all 50 traits are reported in Supplementary Table 11.

**Supplementary Figure 12:**
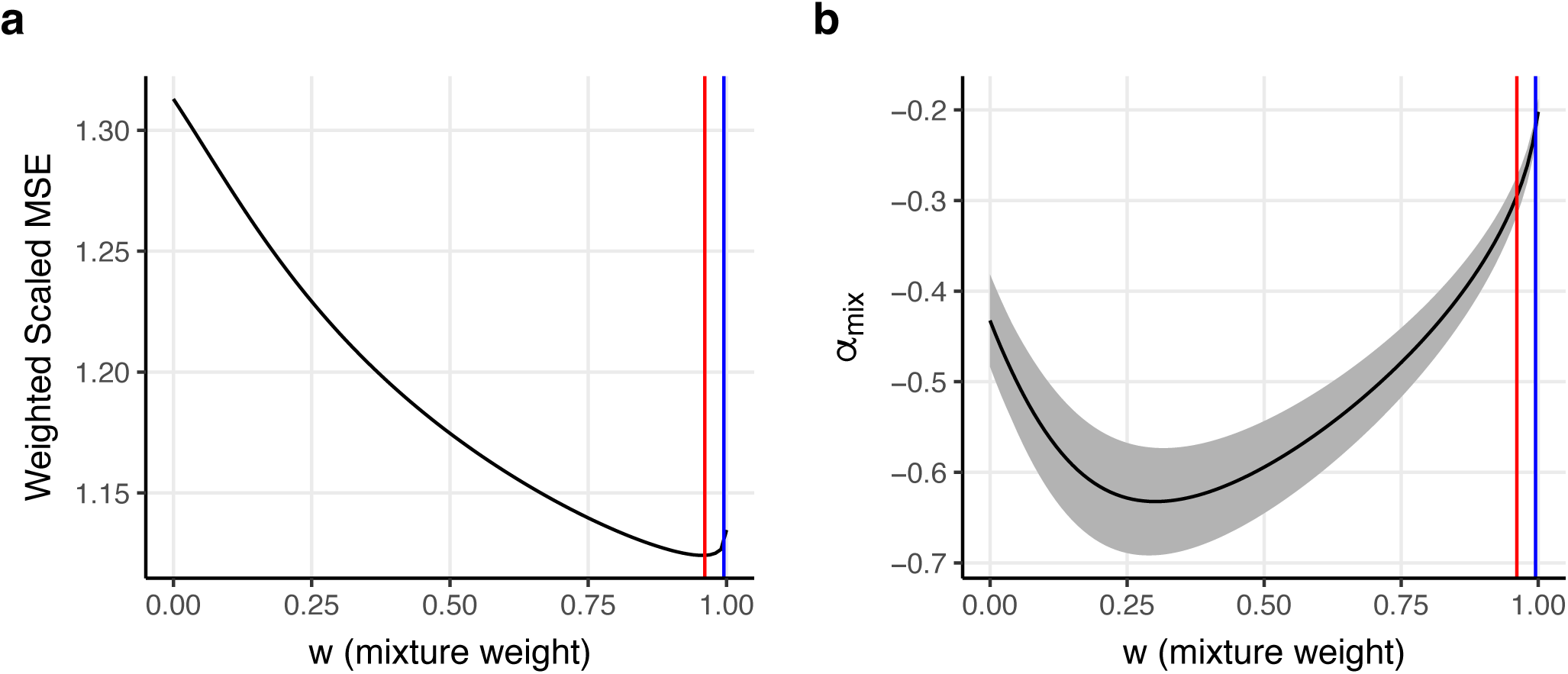
Results of fitting the *α_mix_* model across 50 diseases/traits via a random effects meta-analysis of disease/trait-specific alpha-mix parameters. We report **(a)** the weighted scaled mean squared error, summed across traits, with respect to *w*, and **(b)** estimates of *α_mix_* as a function of *w*. Blue lines denote our meta-analyzed point estimate of *w*. Red lines denote the point estimate of *w* from the main analysis. The shaded region in (b) denotes 95% confidence intervals for *α_mix_*. Weighted scaled MSE was scaled by an arbitrary constant. The random effects meta-analysis estimate of *w* is not guaranteed to minimize the MSE because *w* is estimated separately for each trait, and some traits have MSE functions which are minimized at values other than the minimum of the shared MSE.

**Supplementary Figure 13:**
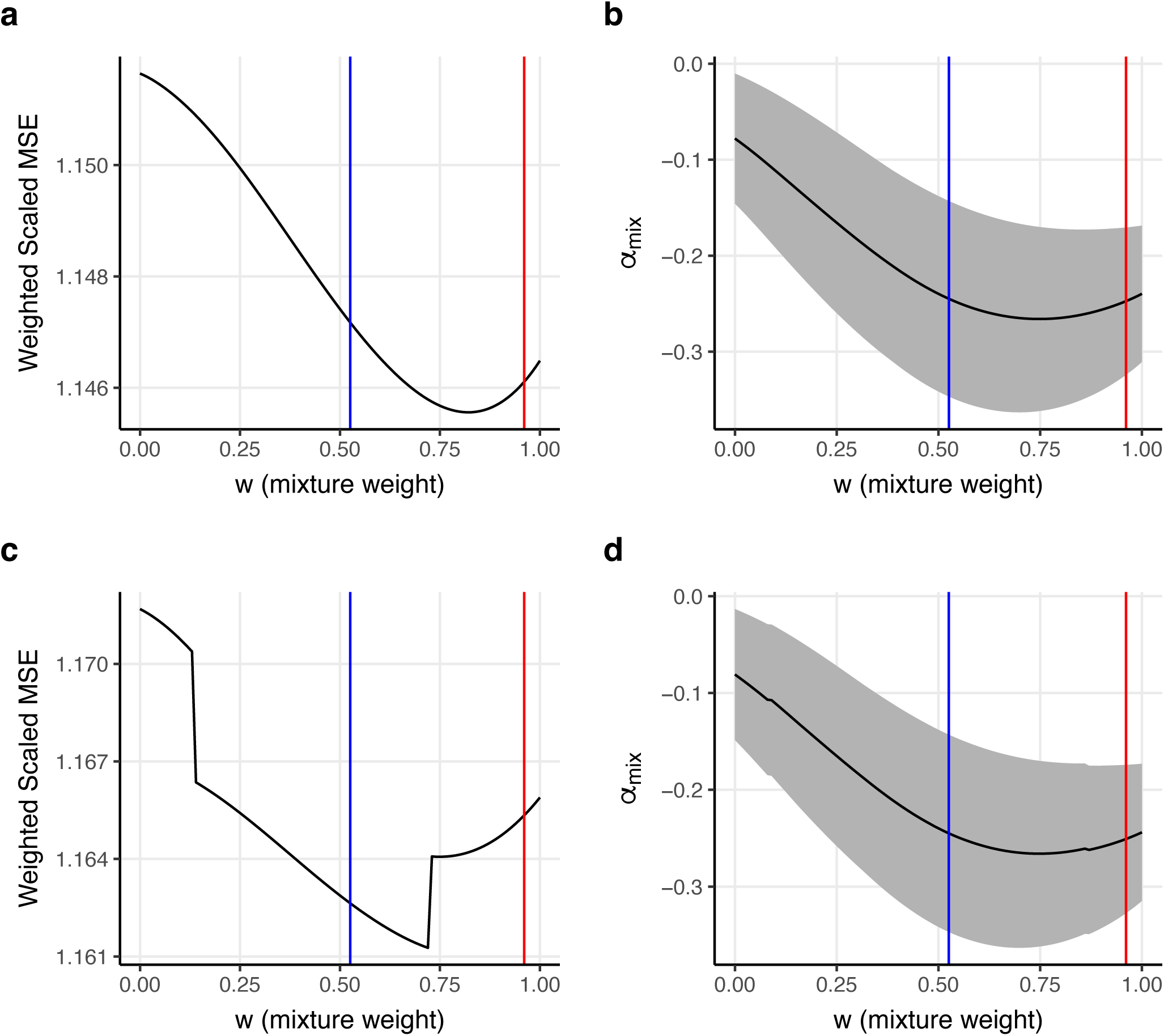
Results of fitting the *α_mix_* model across 50 diseases/traits via a random effects meta-analysis of disease/trait-specific alpha-mix parameters restricted to SNPs with *p_A_* ≥ 0.05 and *p_E_* ≥ 0.05. We report **(a)** the weighted scaled mean squared error with respect to *w*, summed across diseases/traits while restricting to SNPs with *p_A_* ≥ 0.05 and *p_E_* ≥ 0.05, excluding UKB_460K.biochemistry_Phosphate, **(b)** *α_mix_* as a function of *w*, estimated using a random effects meta-analysis across traits while restricting to SNPs with *p_A_* ≥ 0.05 and *p_E_* ≥ 0.05 excluding UKB_460K.biochemistry_Phosphate, **(c)** same as (a), including UKB_460K.biochemistry_Phosphate, and **(d)** same as (b), including UKB_460K.biochemistry_Phosphate. Blue lines denote our meta-analyzed point estimate of *w*. Red lines denote the point estimate of *w* from the main analysis. The shaded regions in (b,d) denote 95% confidence intervals for *α_mix_*. The random effects meta-analysis estimate of *w* is not guaranteed to minimize the MSE because *w* is estimated separately for each trait, and some traits have MSE functions which are minimized at values other than the minimum of the shared MSE function. Inclusion of UKB_460K.biochemistry_Phosphate had very little effect on our meta-analyzed estimates of *w* (difference of .0052) and *α_mix_* (difference of .0040). Weighted scaled MSE was scaled by an arbitrary constant.

**Supplementary Figure 14:**
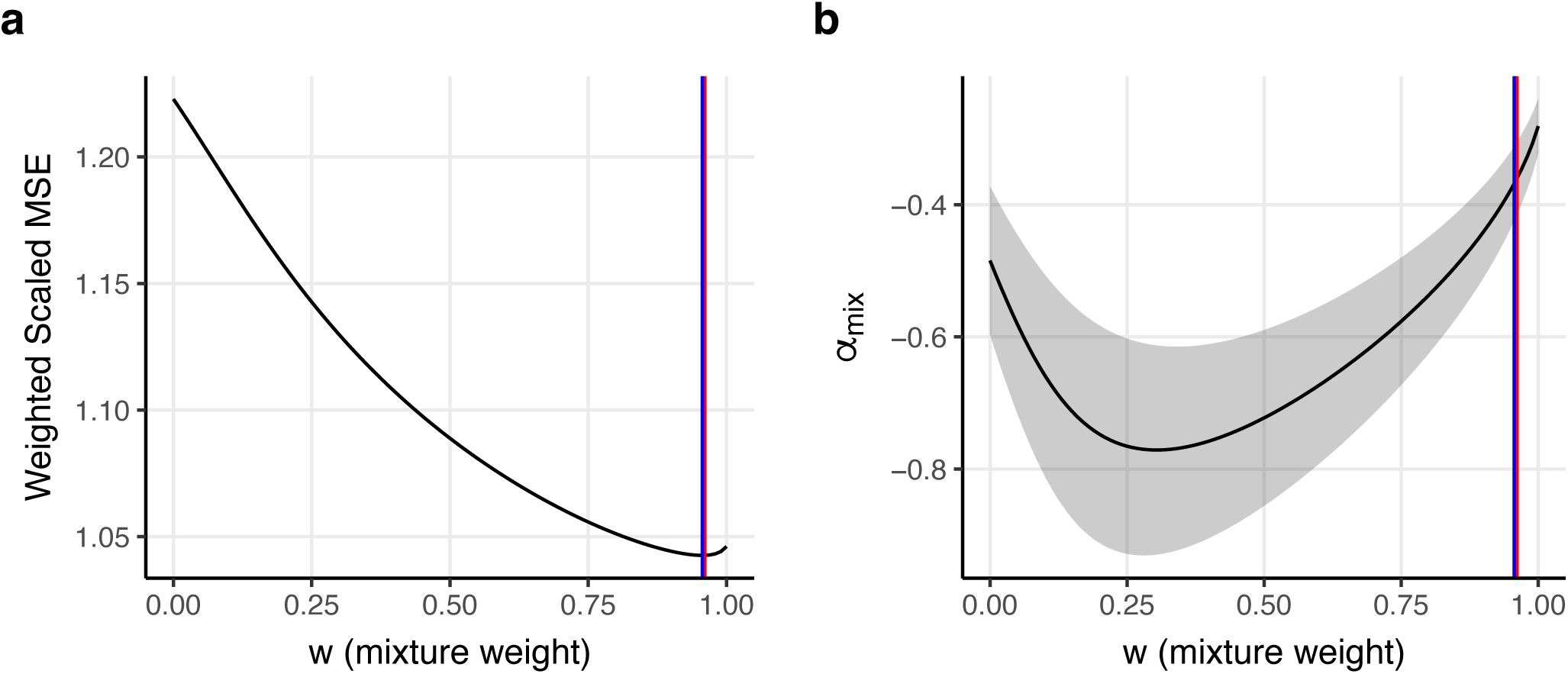
Results of fitting the *α_mix_* model across 50 diseases/traits while estimating African MAF using *N*=107 Yoruban genomes. We report **(a)** the weighted scaled mean squared error with respect to *w*, and **(b)** estimates of *α_mix_* as a function of *w*. Blue lines denote our point estimate of *w* while estimating African MAF using *N*=107 Yoruban genomes. Red lines denote the point estimate of *w* from the main analysis. The shaded region in (b) denotes 95% confidence intervals for *α_mix_*. Yoruban genomes were used to define the 2-dimensional MAF bins which are used both while estimating per-allele effect size variance using S-LDSC and while estimating the parameters of the *α_mix_* model. Weighted scaled MSE was scaled by an arbitrary constant.

**Supplementary Figure 15:**
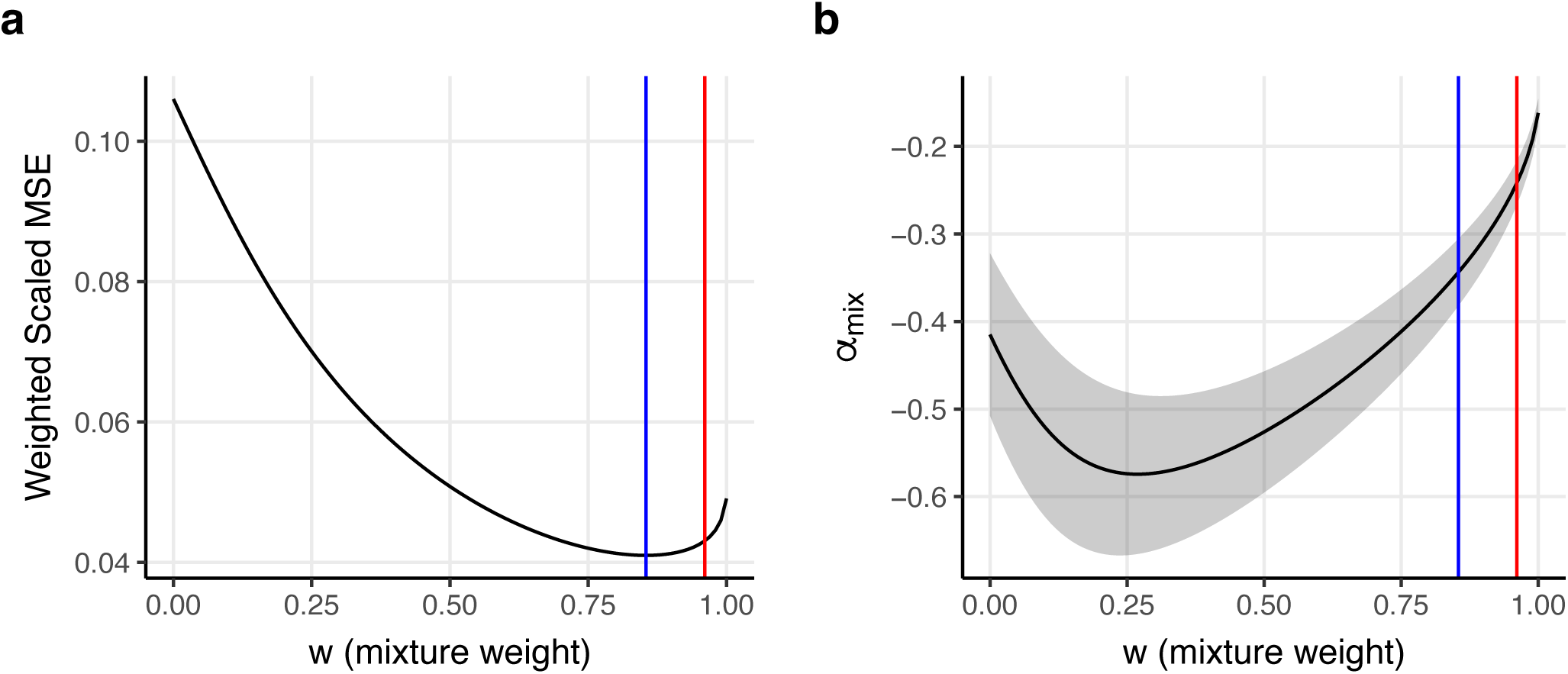
Results of fitting the *α_mix_* model across 50 diseases/traits using the standard Baseline-LD (v2.2) model without annotations for African MAF. We report **(a)** the weighted scaled mean squared error with respect to *w*, and **(b)** estimates of *α_mix_* as a function of *w*. Blue lines denote our point estimate of *w* using the standard Baseline-LD (v2.2) model without annotations for African MAF. Red lines denote the point estimate of *w* from the main analysis. The shaded region in (b) denotes 95% confidence intervals for *α_mix_*. The European MAF bins included in the standard Baseline-LD (v2.2) model were retained. Weighted scaled MSE was scaled by an arbitrary constant.

**Supplementary Figure 16:**
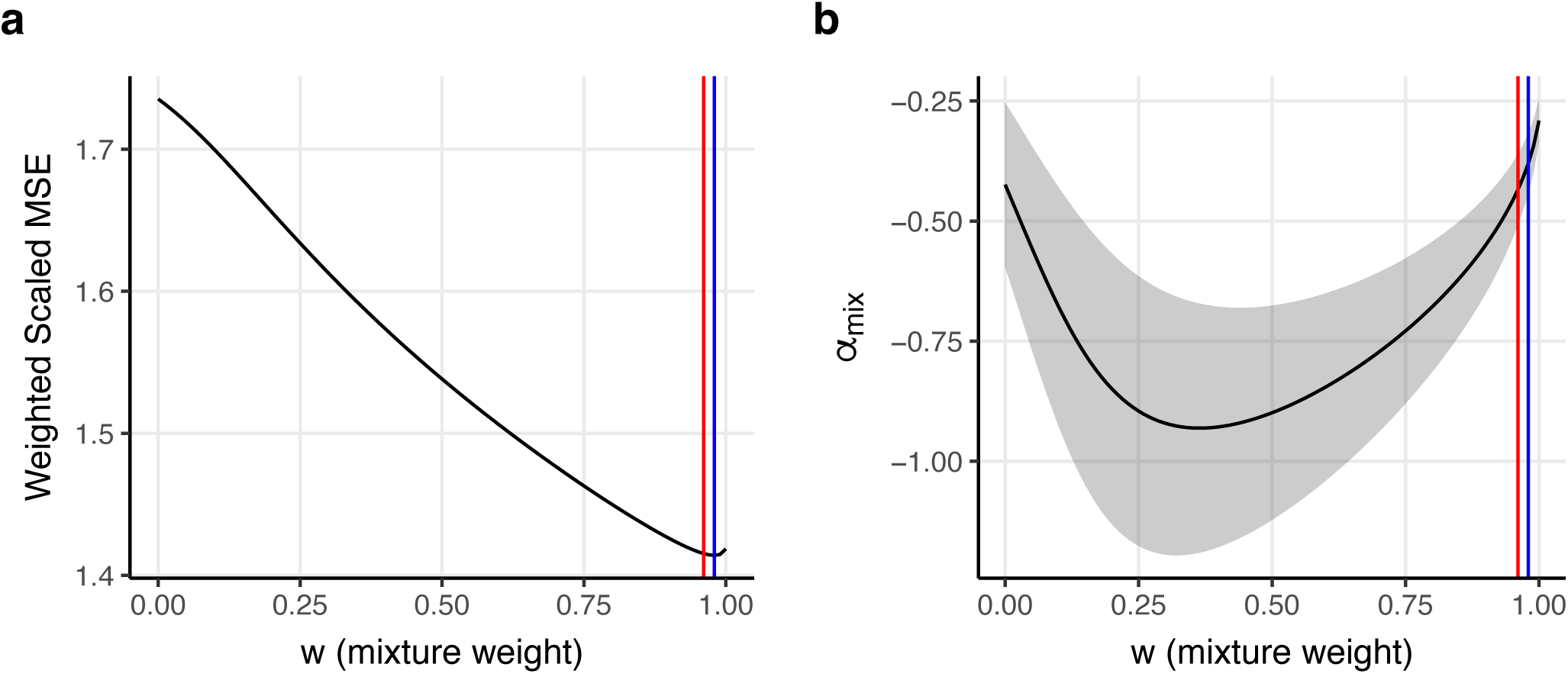
Results of fitting the *α_mix_* model across 50 diseases/traits using 2-dimensional MAF bin annotations without including the BaselineLD-v2.2 model. We report **(a)** the weighted scaled mean squared error with respect to *w*, and **(b)** estimates of *α_mix_* as a function of *w*. Blue lines denote our point estimate of *w* using 2-dimensional MAF bin annotations without including the BaselineLD-v2.2 model. Red lines denote the point estimate of *w* from the main analysis. The shaded region in (b) denotes 95% confidence intervals for *α_mix_*. Weighted scaled MSE was scaled by an arbitrary constant.

**Supplementary Figure 17:**
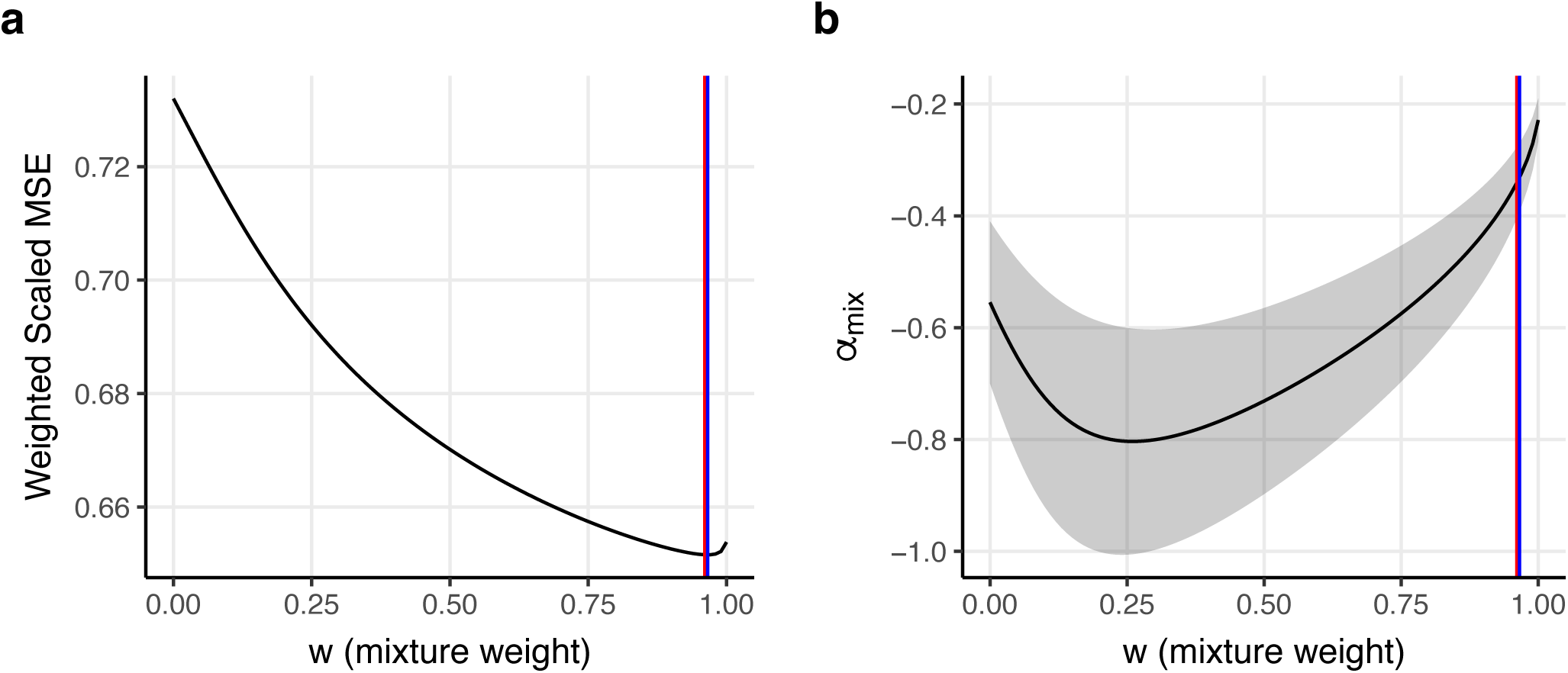
Results of fitting the *α_mix_* model across 29 diseases/traits from UK Biobank. We report **(a)** the weighted scaled mean squared error with respect to *w*, and **(b)** estimates of *α_mix_* as a function of *w*. Blue lines denote our point estimate of *w* using 29 UK Biobank diseases/traits. Red lines denote the point estimate of *w* from the main analysis. The shaded region in (b) denotes 95% confidence intervals for *α_mix_*. Weighted scaled MSE was scaled by an arbitrary constant.

**Supplementary Figure 18:**
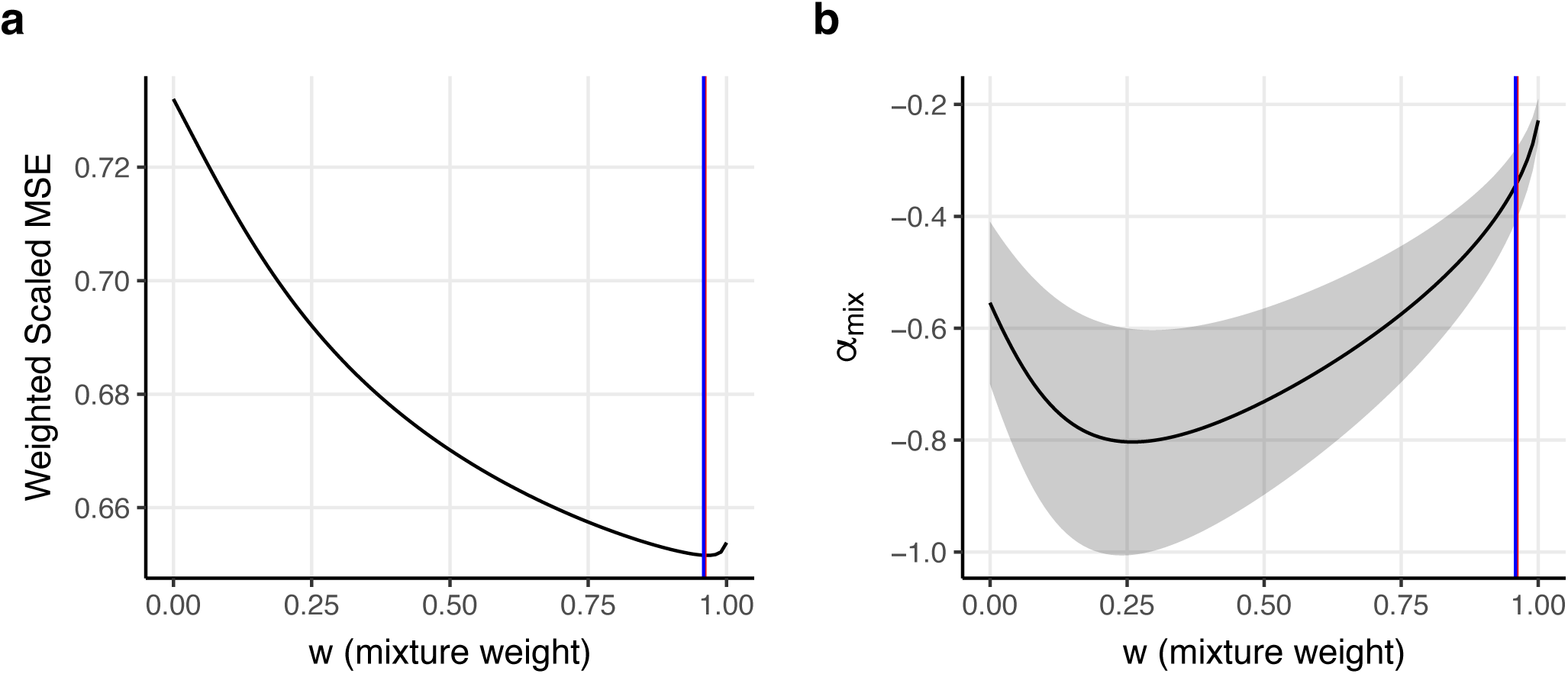
Results of fitting the *α_mix_* model across 21 diseases/traits from publicly available summary statistics. We report **(a)** the weighted scaled mean squared error with respect to *w*, and **(b)** estimates of *α_mix_* as a function of *w*. Blue lines denote our point estimate of *w* using 21 diseases/traits from publicly available summary statistics (not including any diseases/traits from UK Biobank). Red lines denote the point estimate of *w* from the main analysis. The shaded region in (b) denotes 95% confidence intervals for *α_mix_*. Weighted scaled MSE was scaled by an arbitrary constant.

**Supplementary Figure 19:**
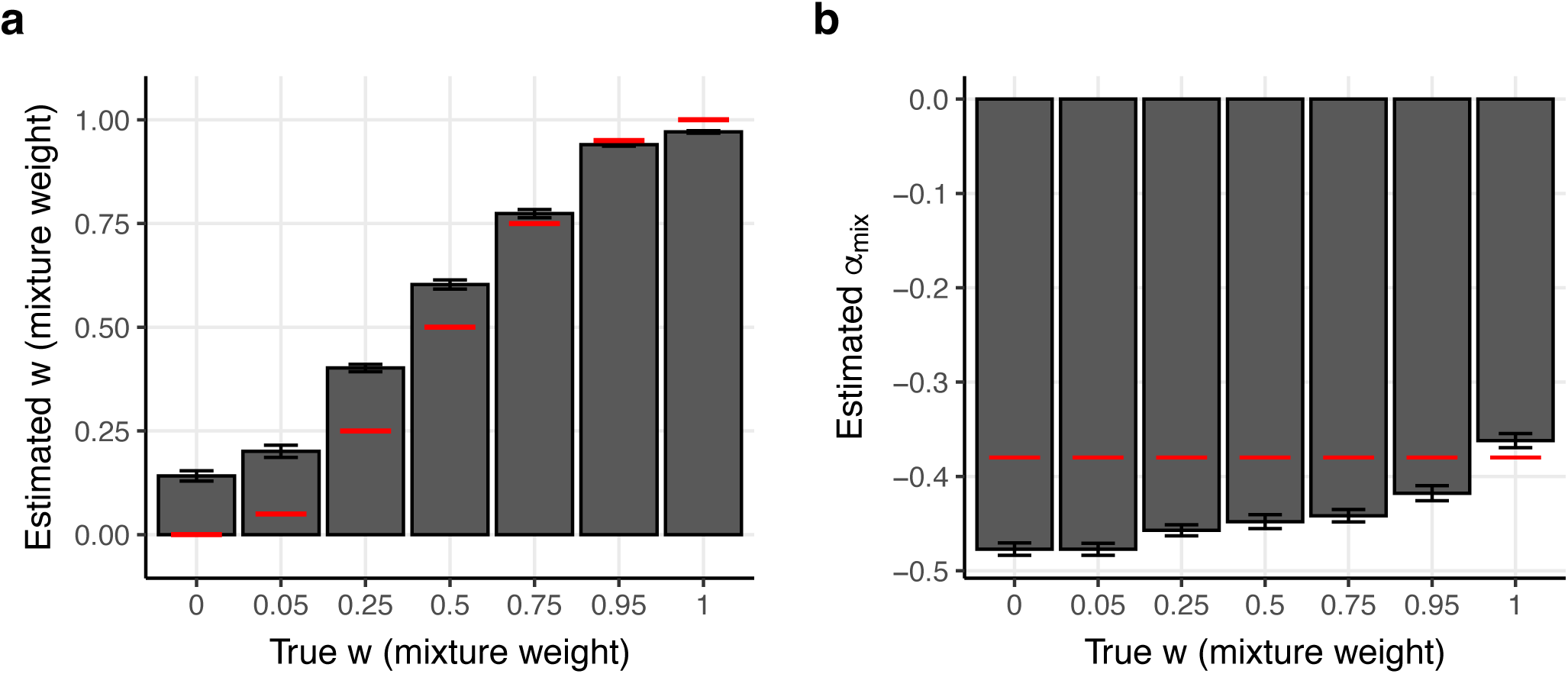
Results of fitting the *α_mix_* model across groups of 25 traits in simulations, while using European MAF from UK Biobank for the generative model, and European MAF from 1000 Genomes for inference. We report estimates of *w* **(a)** and *α_mix_* **(b)**. Bars denote the mean estimate of each parameter across 40 replicates. Red lines denote the true value of each parameter. Error bars denote 95% confidence intervals. 1000 Genomes MAF was used both to define MAF bin annotations given to S-LDSC and during *α_mix_* inference.

**Supplementary Figure 20:**
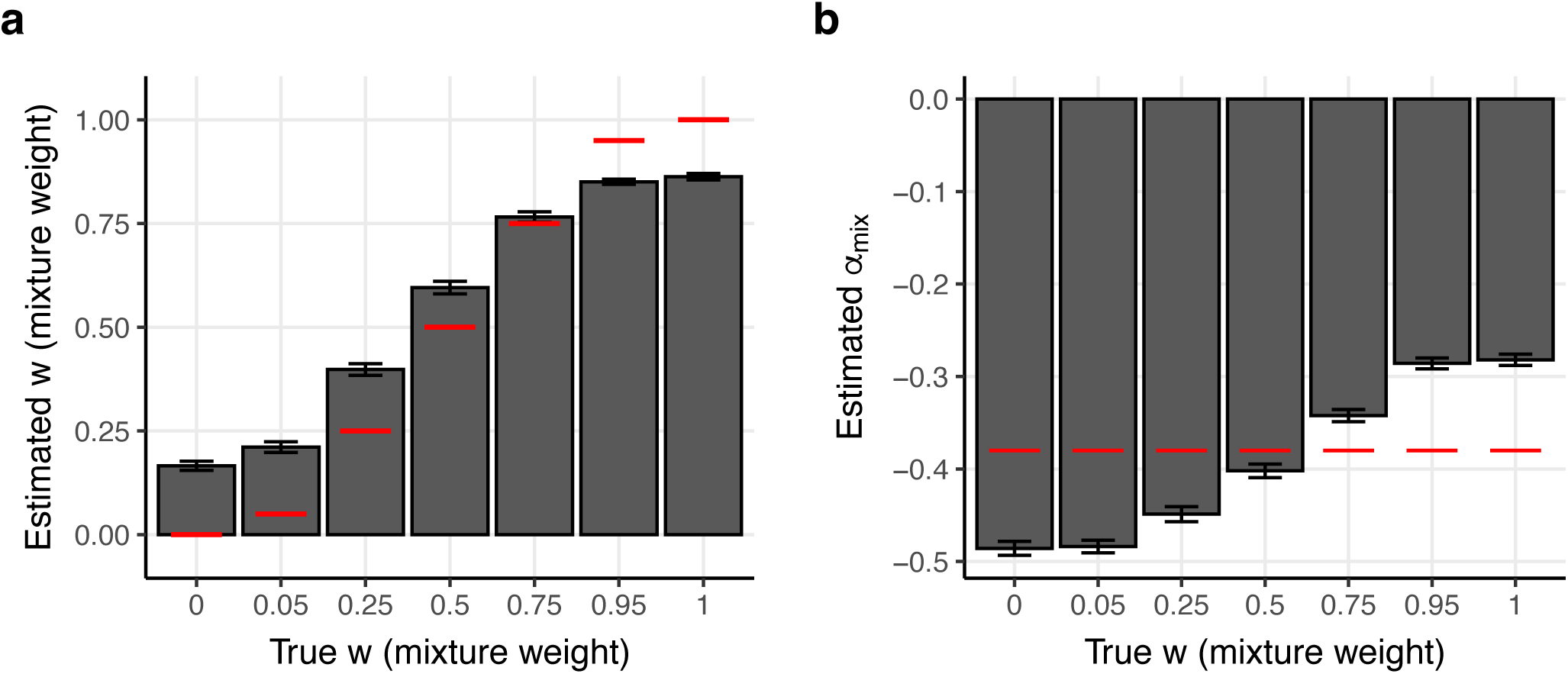
Results of fitting the *α_mix_* model across groups of 25 traits in simulations, while thresholding MAF at *T*=0.05. We report estimates of *w* **(a)** and *α_mix_* **(b)**. Bars denote the mean estimate of each parameter across 40 replicates. Red lines denote the true value of each parameter. Error bars denote 95% confidence intervals. *T* is only used by the generative model, not by S-LDSC or in *α_mix_* inference.

**Supplementary Figure 21:**
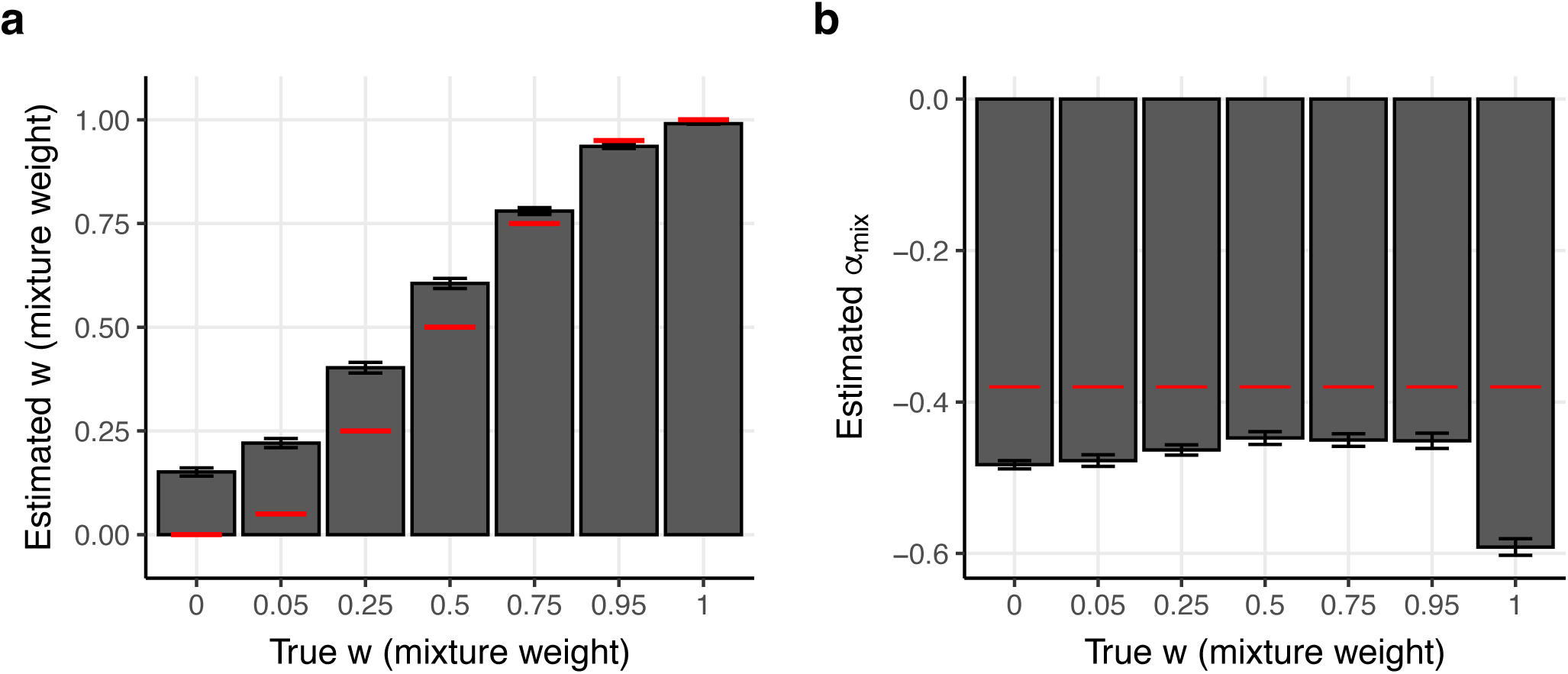
Results of fitting the *α_mix_* model across groups of 25 traits in simulations, while thresholding MAF at *T*=1.5*10^-5^ (equivalent to a minor allele count of 1 in our sample of genomes with local African ancestry from All of Us). We report estimates of *w* **(a)** and *α_mix_* **(b)**. Bars denote the mean estimate of each parameter across 40 replicates. Red lines denote the true value of each parameter. Error bars denote 95% confidence intervals. *T* is only used by the generative model, not by S-LDSC or in *α_mix_* inference.

**Supplementary Figure 22:**
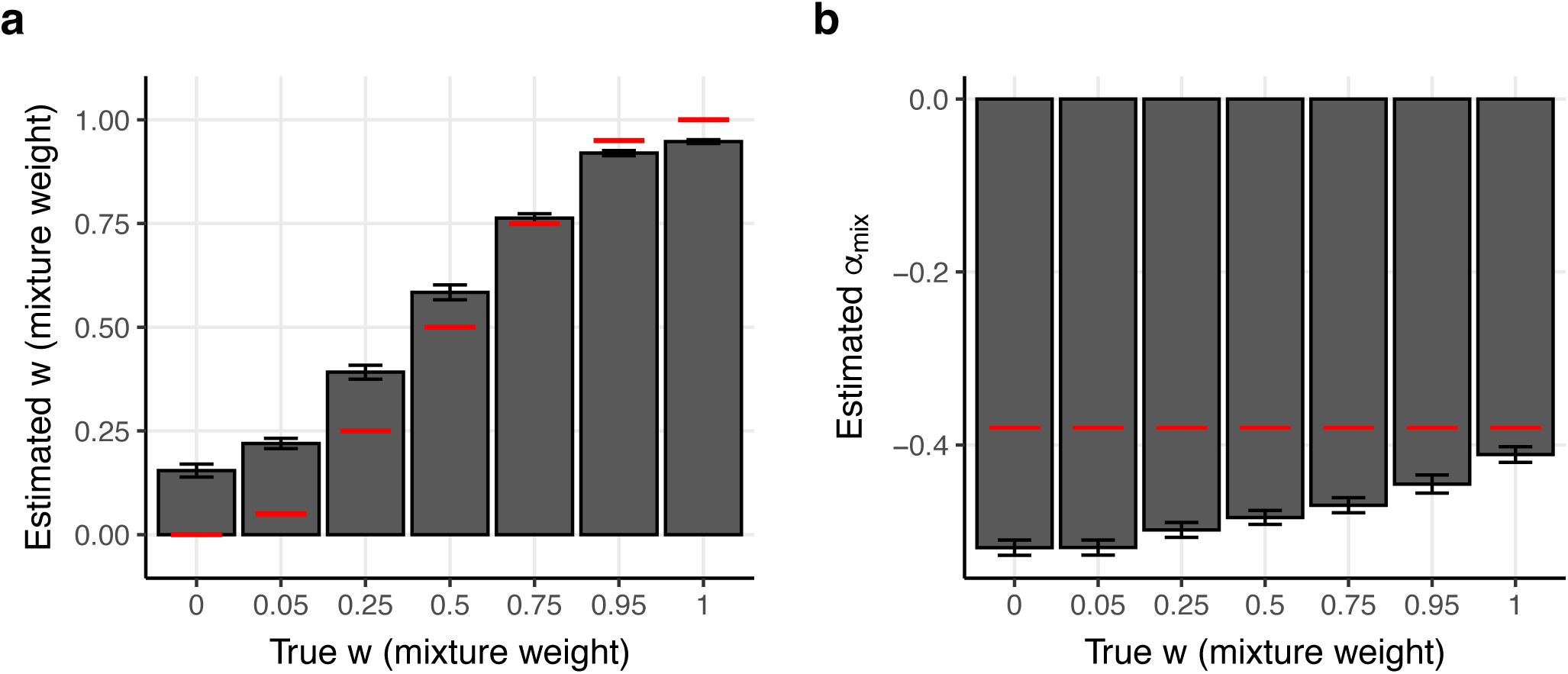
Results of fitting the *α_mix_* model across groups of 25 traits in simulations for traits with 5,000 causal variants. We report estimates of *w* **(a)** and *α_mix_* **(b)**. Bars denote the mean estimate of each parameter across 40 replicates. Red lines denote the true value of each parameter. Error bars denote 95% confidence intervals.

**Supplementary Figure 23:**
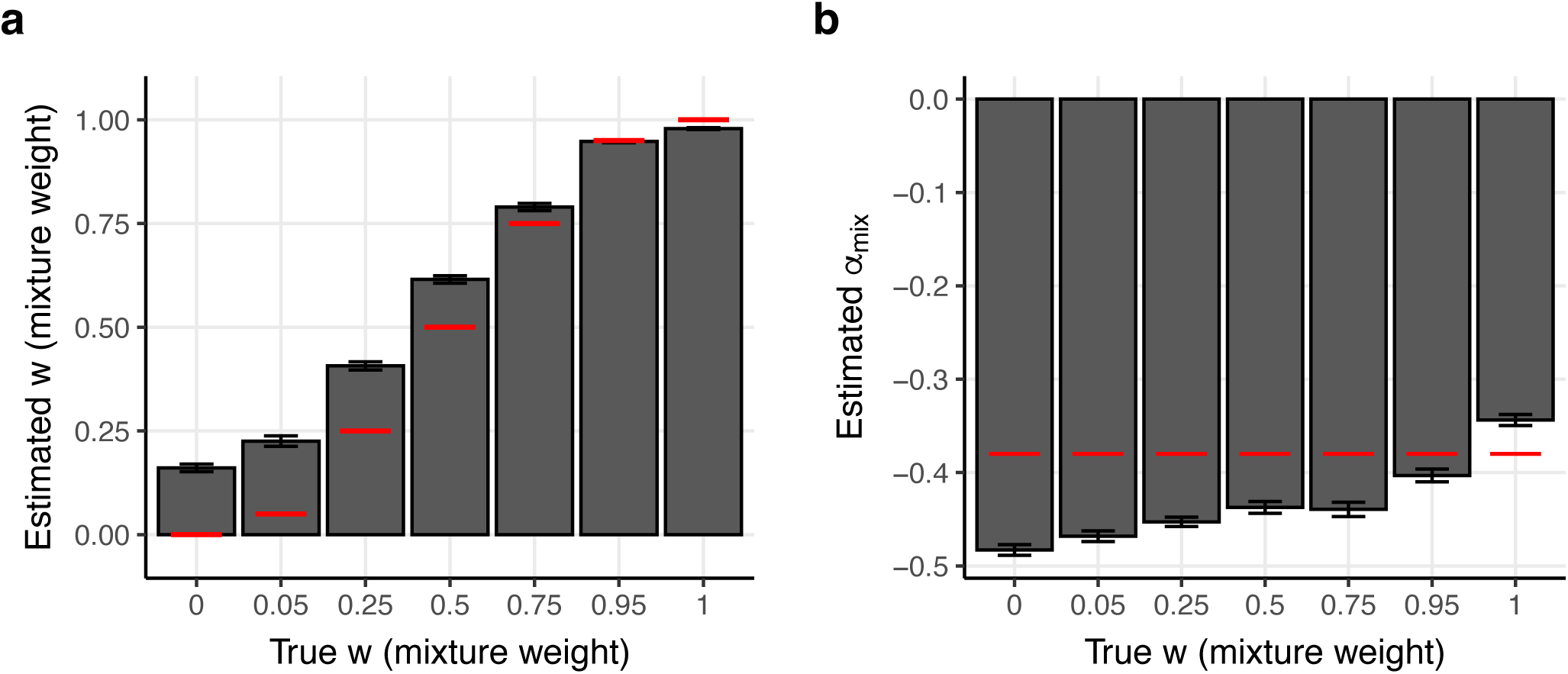
Results of fitting the *α_mix_* model across groups of 25 traits in simulations for traits with 20,000 causal variants. We report estimates of *w* **(a)** and *α_mix_* **(b)**. Bars denote the mean estimate of each parameter across 40 replicates. Red lines denote the true value of each parameter. Error bars denote 95% confidence intervals.

**Supplementary Figure 24:**
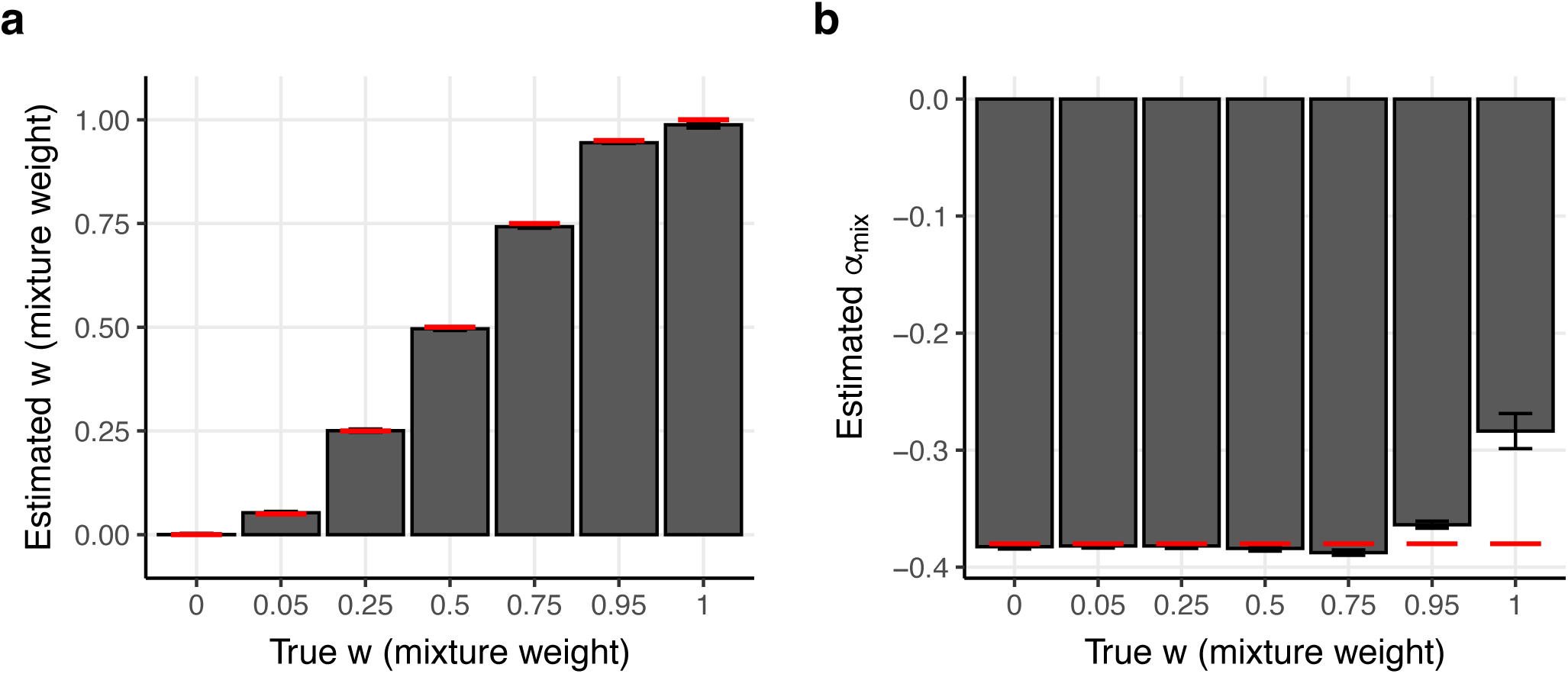
Results of fitting the *α_mix_* model across groups of 25 traits in simulations using exact values of *β*^2^ during inference, rather than estimates from S-LDSC. We report estimates of *w* **(a)** and *α_mix_* **(b)**. Bars denote the mean estimate of each parameter across 40 replicates. Red lines denote the true value of each parameter. Error bars denote 95% confidence intervals.

**Supplementary Figure 25:**
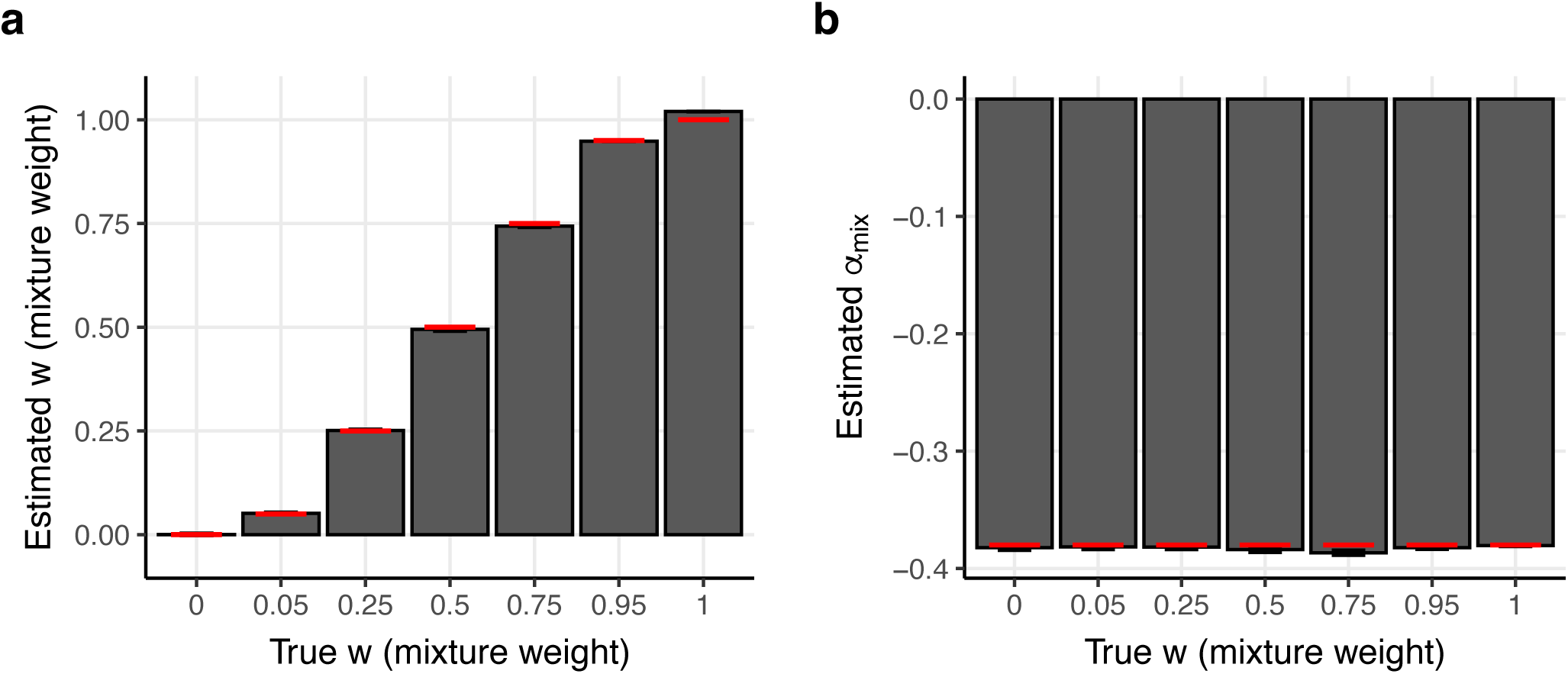
Results of fitting the *α_mix_* model across groups of 25 traits in simulations while using exact values of *β*^2^ (rather than estimates from S-LDSC), values of *p_A_* and *p_E_* derived from causal variants (rather than all variants), and thresholding *p_mix_* at *T*=0.005. We report estimates of *w* **(a)** and *α_mix_* **(b)**. Bars denote the mean estimate of each parameter across 40 replicates. Red lines denote the true value of each parameter. Error bars denote 95% confidence intervals. *p_mix_* was computed separately for each causal SNP, thresholded at *T*=0.005, and then averaged over all causal SNPs in each two-dimensional MAF bin.

**Supplementary Figure 26:**
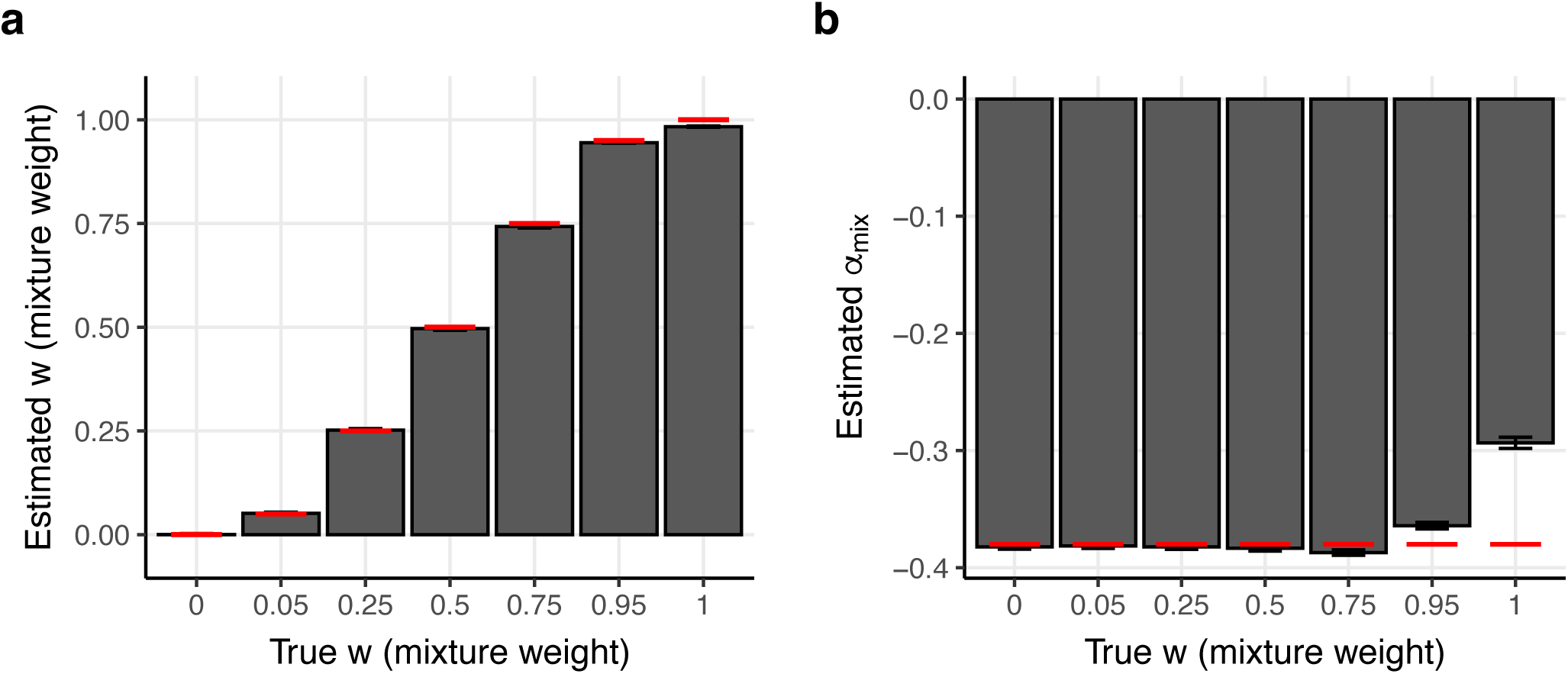
Results of fitting the *α_mix_* model across groups of 25 traits in simulations while using exact values of *β*^2^ (rather than estimates from S-LDSC) and values of *p_A_* and *p_E_* derived from causal variants (rather than all variants). We report estimates of *w* **(a)** and *α_mix_* **(b)**. Bars denote the mean estimate of each parameter across 40 replicates. Red lines denote the true value of each parameter. Error bars denote 95% confidence intervals. *p_mix_* was computed separately for each causal SNP, thresholded at 1.5*10^-5^ (equivalent to a minor allele count of 1 in our sample of genomes with local African ancestry from All of Us), and then averaged over all causal SNPs in each two-dimensional MAF bin.

**Supplementary Figure 27:**
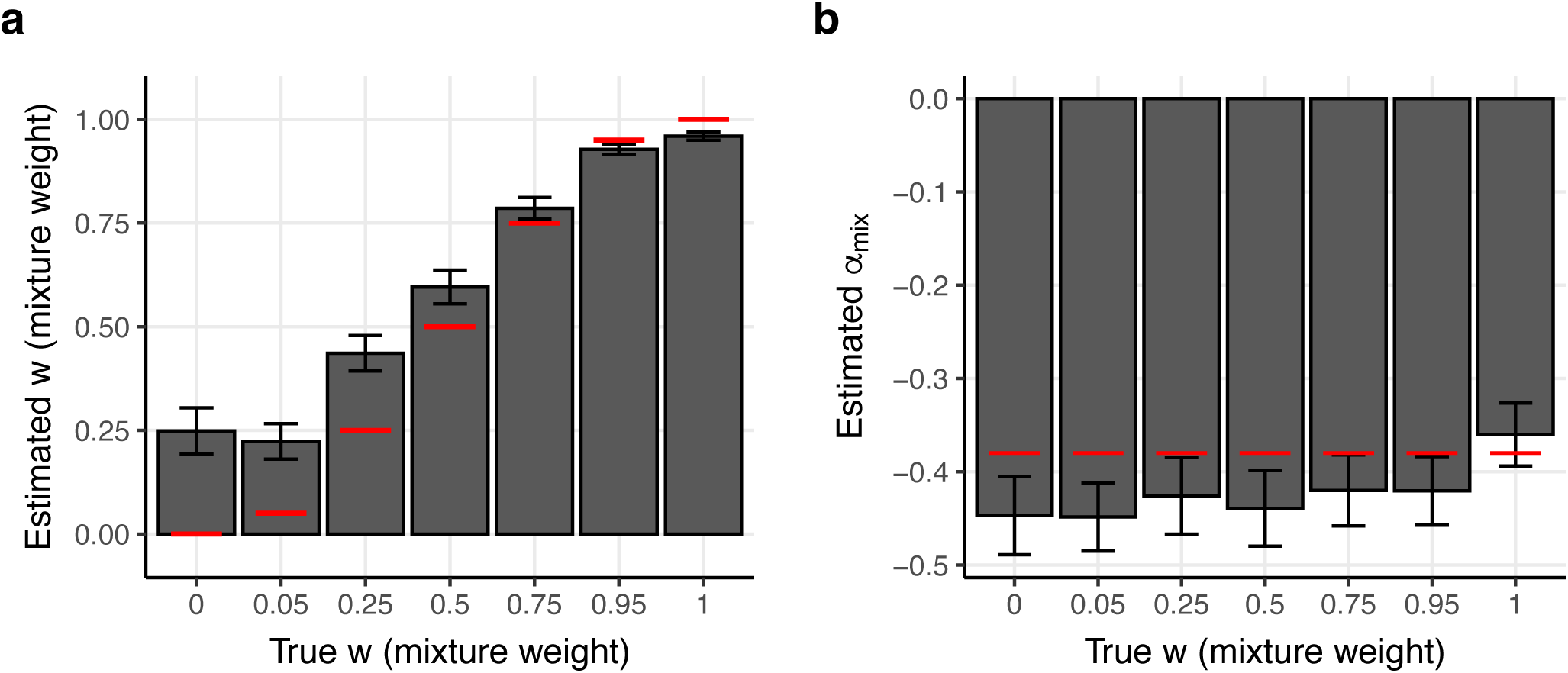
Results of fitting the *α_mix_* model separately for each trait in simulations. We report estimates of *w* **(a)** and *α_mix_* **(b)**. Bars denote the mean estimate of each parameter across 100 replicates (each with a single trait). Red lines denote the true value of each parameter. Error bars denote 95% confidence intervals.

## Methods

### Minor allele frequency estimation

Throughout this manuscript we refer to unadmixed African MAF as African MAF. We used continental ancestry group assignments provided by All of Us^42^ to identify individuals residing in the United States with predominantly African ancestry; we note that segments of African ancestry in these individuals are primarily of West African origin^48^, such that our African MAF estimates primarily represent West African ancestry. We defined the MAF of a SNP in a specific ancestry to be the frequency of the less common (minor) allele *in that ancestry*. As a result, for a single SNP, it is possible for the minor allele in Africans to be different from the minor allele in Europeans.

We estimated unadmixed African MAF using All of Us^42^ (see Data Availability) whole genome-sequencing data for *N*=46,672 unrelated individuals labeled as African ancestry by All of Us (AoU). This cohort has moderate levels of European admixture on average, so estimating unadmixed allele frequencies (AF) is more complex than in an unadmixed cohort^74^. We restricted to 30,906,411 single-nucleotide polymorphisms with either European MAF ≥ 0.001 or admixed African MAF ≥ 0.001 (as reported by the AoU Variant Annotation Table) in the All of Us v8 short read whole-genome sequencing Allele Count/Allele Frequency threshold Plink dataset. We first estimated the local ancestry of each individual using RFMix2 ^49^ with 1000 Genomes^44^ YRI, CEU, PEL, EAS (concatenation of CHS, CDX, KHV, CHB, JPT), and SAS (concatenation of PJL, BEB, STU, ITU, GIH) as the reference populations, restricting to HapMap3 variants. The local ancestry of SNPs that were not included in HapMap3 was interpolated between that of the closest HapMap3 SNPs. Next, we computed the unadmixed African allele frequency of each SNP using Plinkv1.9^75^ while restricting to individuals with two African haplotypes at that SNP.

We estimated unadmixed European MAF in 70,346 unrelated European ancestry individuals from the All of Us v8 short read whole-genome sequencing Allele Count/Allele Frequency threshold Plink dataset, restricting to the same set of variants used for African MAF estimation. These European MAF estimates were used for analyses of non-synonymous variation (Figures 1-2), but not analyses for analyses of diseases/traits (Figures 3-6).

For S-LDSC analyses of diseases/traits, unadmixed African AF was estimated as described above, but using the All of Us v7.1 short read whole-genome sequencing Allele Count/Allele Frequency threshold Plink dataset. AF were converted from GRCh38 to GRCh37 using BCFtools/liftover^76^. The African MAF of SNPs in 1000 Genomes that could not be matched to any of the SNPs from AoU, was calculated using *N*=107 individuals from Yoruba in 1000 Genomes restricting to unrelated, QC’ed individuals from TGP2261 from ref.^77^. For S-LDSC analyses of diseases/traits, European AF was calculated using *N*=489 European ancestry individuals from 1000 Genomes restricting to unrelated, QC’ed individuals from TGP2261 from ref.^77^. We used 1000 Genomes rather than the All of Us to ensure consistency between our MAF estimates and those used in the Baseline-LD (v2.2) model, where MAF and LD scores are calculated using the same 489 individuals from 1000 Genomes.

Throughout this manuscript, we stratified variants by African and European allele frequency using a common set of bin boundaries, which are justified as follows. SNPs were first stratified by common (MAF ≥ 0.05), low-frequency (0.005 ≤ MAF < 0.05), and rare (MAF < 0.005) categories. In some analyses, we further stratified common variants into 5 or 10 bins with boundaries corresponding to 1000 Genomes European ancestry common MAF quintiles or deciles as defined by the Baseline-LD (v2.2) model. We used 5 common variant bins for visualization (e.g. Figures 1a, 3a) and in-text reporting of non-parametric MAF-dependence statistics (paragraph 1 of the *Per-allele disease and complex trait effect sizes are predominantly African MAF-dependent in European populations* subsection of Results) to improve interpretability. We used 10 common variant bins when fitting the *α_mix_* model (Figures 4-5) because using more bins decreases the absolute difference between the MAF of each SNP in a bin and the mean MAF of the bin. In All of Us analyses, rare SNPs were sometimes further stratified using a boundary of 0.002 to create a rare SNP bin that avoids ascertainment bias (e.g. a SNP with unadmixed African MAF=0.0011 has African American MAF around 0.0008 < 0.001 and thus would typically be excluded by our initial MAF filter; however, a SNP with unadmixed African MAF=0.002 has African American MAF around 0.0016 > 0.001 and thus would typically be included in our dataset).

### Modeling non-synonymous status as a function of MAF

We defined a set of non-synonymous SNPs in AoU by grouping the following values for the “consequence” field in the All of Us v8 Variant Annotation Table: frameshift_variant, inframe_deletion, inframe_insertion, missense_variant, start_lost, stop_gained, stop_lost. We computed the proportion of SNPs that are non-synonymous (PNS) in a stratum with respect to all SNPs, including SNPs in non-coding regions.

We modeled the probability that a SNP is non-synonymous as a function of *p_mix_* via the following logistic regression model:

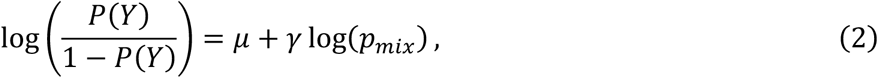

where *Y*=1 if the SNP is non-synonymous and 0 otherwise and *p_mix_*=*w*\**p_A_*+(*1*-*w*)\**p_E_*. If *w*=0 or 1, this is equivalent to fitting a logistic regression using European or African, respectively. To estimate *w*, we computed a profile likelihood, treating 𝜇 and 𝛾 as nuisance parameters, and used a numerical optimization method to maximize the profile likelihood with respect to *w.* The profile likelihood is given by:

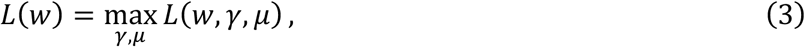

where 𝐿(𝑤, 𝛾, 𝜇) is the likelihood from equation 2. We calculated 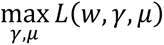 for a specific choice of *w* using discrete.discrete_model.Logit from the statsmodels^78^ Python package and numerically identified the maximum of 𝐿(𝑤) with respect to *w* using optimize.minimize_scalar from the SciPy Python package^79^. While fitting logistic regression models, values of *p_A_* and *p_E_* < 0.002 were set to 0.002, except for in Supplementary Figure 8, where a threshold of 0.05 is used.

Significance testing related to logistic regressions was performed in four ways depending on the specific analysis. We estimated P-values for 𝛾 in *p_A_*-only and *p_E_*-only univariate logistic regressions using a two-tailed t-test. When comparing model fit for a *p_A_*-only model to a *p_E_*-only model on a shared set of SNPs, we computed p-values using Vuong’s test because both models had the same number of degrees of freedom (precluding the use of a standard likelihood ratio test). When comparing *r^2^* and regression coefficients for a *p_A_*-only model to a *p_E_*-only model which were fit on different sets of SNPs, we computed p-values using a genomic block jackknife with 200 approximately equally sized blocks. All *p_mix_* logistic regression *P*-values and confidence intervals were computed by using a likelihood ratio test statistic with 1 degree of freedom. Throughout the manuscript, *P*-values less than 10^-15^ were reported as *P*<10^-15^.

### Disease/trait selection

We first collected a set of 364 GWAS summary statistics in European ancestry cohorts across a wide range of complex traits and diseases. For each complex trait or disease, we estimated heritability using S-LDSC with baseline-LD (v2.2) LD scores. For each pair of complex traits and diseases, we estimated genetic correlation using S-LDSC^80^ without partitioning based on functional annotations. Next, we filtered to traits with heritability z-score greater than 6 and *N*h^2^*>20,000. Finally, we greedily selected traits based on *N*h^2^*, subject to the constraint that for each pair of selected traits, the squared genetic correlation < 0.1. The output of this procedure was 50 diseases/complex traits (average *N*=483K).

### Estimation of per-allele effect size variance via stratified linkage disequilibrium score regression

Stratified linkage disequilibrium score regression^3^ (S-LDSC) estimates the common SNP heritability of a GWAS complex trait or disease partitioned across predefined functional annotations. In S-LDSC, the per-SNP heritability of SNP *i* is modeled as 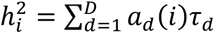, where *d* indexes a set of *D* functional annotations, 𝑎_𝑑_(𝑖) is the value of annotation *d* for SNP *i*, and 𝜏_𝑑_represents the increase in 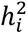 associated with a unit increase in 𝑎_𝑑_(𝑖). S-LDSC requires the specification of a set of *D* functional annotations that explain how heritability varies across the genome.

We applied S-LDSC using annotations from a new extension of the Baseline-LD (v2.2) model^5^. The Baseline-LD (v2.2) model includes standard functional annotations in addition to MAF and linkage disequilibrium (LD) annotations that reduce MAF and LD-related bias. Our extension of the Baseline-LD (v2.2) model incorporated new annotations based on bivariate MAF bins, where each bivariate MAF bin is defined by an African MAF bin and a European MAF bin. Specifically, we defined 12 univariate unadmixed African MAF bins: 10 for common African variants (*p_A_* ≥ 0.05), 1 for low-frequency African variants (0.005 ≤ *p_A_* < 0.05), and 1 for rare African variants (*p_A_* < 0.005). The boundaries of the 10 common variant bins were the same as those used for univariate European common MAF in the Baseline-LD (v2.2) model. We then created 120 bivariate MAF bins by stratifying SNPs based on the interaction between the 12 univariate African MAF bins and 10 univariate European MAF bins. We removed the 10 univariate European MAF bins from the combined model to avoid collinearity of functional annotations. We calculated partitioned LD scores using 489 European ancestry individuals from 1000 Genomes.

To estimate the *per-allele* effect size variance of variant *i*, we divided 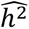 (as estimated by S-LDSC) by the sample variance under Hardy-Weinberg equilibrium, 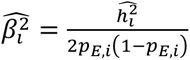, where *p_E,i_* is the European MAF for SNP *i*. This procedure has been previously applied in ref.^81^. We calculated the per-allele effect size variance for a specific trait and set of SNPs, *C*, as 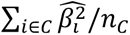, where *n_C_* is the number of SNPs contained in *C*. When meta-analyzing across traits, we normalized 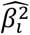 within each trait before taking the mean (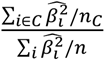, where *n* is the number of SNPs with *p_E_* > 0.05), and meta-analyzed these normalized quantities using random-effect meta-analysis as implemented in stats.meta_analysis.combine_effects from the statsmodels Python package.

### Estimation of alpha-mix parameters

We propose the *α_mix_* model which relates per-allele effect size variance to unadmixed African and European MAF:

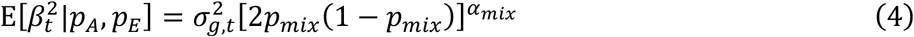

where 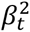 is the squared *per-allele* effect size of a single-nucleotide polymorphism (SNP) in trait *t*, 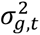 is a variance parameter, *p_mix_*=*w*\**p_A_ + (1-w)*p_E_*, *w* specifies the relative contributions of *p_A_* and *p_E_*, and *α_mix_* specifies the level of MAF dependence. Equation 4 is identical to equation 1 with a trait-specific subscript.

We estimated the parameters of the *α_mix_* model via a generalized method of moments estimator, with population moments from equation 4 and conditional sample moments estimated via S-LDSC. We first partitioned variants with European MAF ≥ 0.05 into 120 two-dimensional MAF bins based on the interaction between 12 African MAF bins and 10 European MAF bins. We then defined one moment condition for each trait-bin pair: 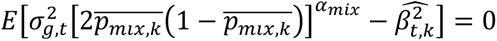 where 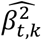, is the S-LDSC estimate of the per-allele effect size variance (described in *Estimation of per-allele effect size variance via stratified linkage disequilibrium score regression*) and 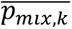 is the mean of *p_mix_* across all SNPs in bin *k*.

We next applied a numerical optimization procedure to minimize the sum of squared moment conditions, weighted by the number of SNPs in each bin. When estimating alpha-mix parameters for a single trait, we optimized the following loss function: 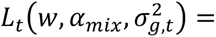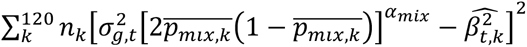, using the differential evolution optimization method with default parameters as implemented by the SciPy package to search over *α_mix_* ∈ [-1.5, 1.5] and 𝑤 ∈ [-0.5,1.5], while solving for 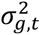 analytically (see below).

When estimating alpha-mix parameters jointly across *T* traits, we optimized the following loss function:

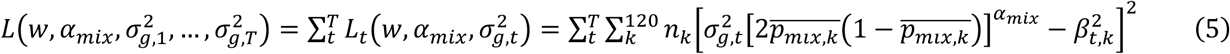

To avoid optimizing this function numerically with respect to *T* + 2 unknown parameters, note that for a fixed choice of *w* and *α_mix_*,

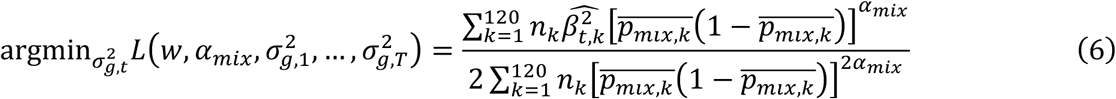

so we can treat this as an optimization problem with two unknown parameters, *w* and *α_mix_*, and use equation 6 to identify 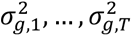 analytically, in turn, for each trait. We minimized *w* and *α_mix_*using the SciPy package implementation of the differential evolution method with default parameters, restricting *α_mix_* to [-1.5, 1.5], and 𝑤 to [-0.5, 1.5].

To estimate uncertainty of parameter estimates and perform significance testing, we wrapped the entire procedure in a genomic block-jackknife with 200 blocks (including both S-LDSC parameter estimation and GMM estimation). When estimating *α_mix_* and *w* jointly across traits, we used a modified version of S-LDSC which forces jackknife block boundaries to be identical across summary statistics that include distinct sets of SNPs.

### Simulations

We simulated quantitative traits using imputed genotypes for 337,448 unrelated British individuals from UK Biobank (version 3 of the Haplotype Reference Consortium imputed data) and 5,907,305 autosomal SNPs with *p_E_* ≥ 0.05 in 1000 Genomes and INFO score ≥ 0.60. Quantitative traits were simulated under a standard additive model: 𝑌 = 𝑋𝛽 +ε, where 𝑌 is a phenotype vector, 𝑋 is a genotype matrix, 𝛽 is a sparse vector with 𝑛_𝑐𝑎𝑢𝑠𝑎𝑙_ non-zero entries representing per-allele causal effects, and 𝜖 is normally distributed residual variance. The non-zero elements of 𝛽 were sampled separately for each trait. The effects of non-zero elements of 𝛽 were sampled from a normal distribution with mean 0 and variance 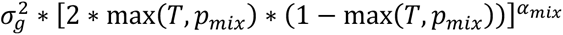 where 𝑇 represents a threshold below which varying MAF does not change effect size (see left side of Figure 2 from ref.^19^). When simulating effects, we used 1000 Genomes for European MAF, and All of Us and 1000 Genomes allele frequencies for African MAF (see the *Minor allele frequency estimation* subsection of Methods), except when stated otherwise. 𝛽 was scaled such that 𝑣𝑎𝑟(𝑋𝛽) = 𝑣𝑎𝑟(𝑌)/2 to set *h^2^*=0.5. GWAS and computation of 𝑋𝛽 were performed using Plink v2.0^82^.

## Code Availability

We have publicly released custom code used to estimate *α_mix_* and perform all analyses described in this manuscript at https://github.com/jordanero/alphamix.

## Data Availability

GWAS summary statistics and S-LDSC results for all diseases/traits are publicly available at https://zenodo.org/records/18292998. Unadmixed African MAF estimates are available at https://zenodo.org/records/18292998 (MAF corresponding to a minor allele count less than 40 has been censored in accordance with the All of Us Data and Statistics Dissemination Policy). AoU short read individual-level WGS data are available to authorized users on the All of Us Researcher Workbench. The UKBB resource is publicly available by application (http://www.ukbiobank.ac.uk).

## Acknowledgements

We are grateful to Xihong Lin, Po-Ru Loh, David Reich, Xilin Jiang, Mashaal Sohail, Bogdan Pasaniuc, Sasha Gusev, and Benedikt Geiger for helpful discussions. We thank All of Us participants for making this research possible. We also thank All of Us for providing access to the data used in this research. This research was conducted using the UK Biobank resource under application no. 16549. This research was funded by National Institutes of Health (NIH) grants R01 MH101244, R37 MH107649, R01 HG006399, F31 HG013040, and T32 GM135117. The funders had no role in study design, data collection and analysis, decision to publish or preparation of the manuscript.

## Competing interests

J.R. is an employee of Genentech. The remaining authors declare no competing interests.

## Notes

### Funding Statement

This research was funded by National Institutes of Health (NIH) grants R01 MH101244, R37 MH107649, R01 HG006399, U01 HG012009, F31 HG013040, and T32 GM135117.

### Author Declarations

All of Us whole-genome sequencing data is available to authorized users on the AoU Researcher Workbench (https://workbench.researchallofus.org/) (data was available prior to study initiation). UK Biobank data is available at http://www.ukbiobank.ac.uk (data was available prior to study initiation).

### Summary of Updates

Added additional simulation secondary analyses and made small text changes.

